# Plasma and CSF proteomic signatures related to Alzheimer’s, α-synuclein, or vascular pathologies and clinical decline

**DOI:** 10.64898/2026.02.04.26345534

**Authors:** Anna Orduña Dolado, Alexa Pichet Binette, Andréa L. Benedet, Ilaria Pola, Kübra Tan, Wiebke Traichel, Ines Hristovska, Angela Mammana, Erik Stomrud, Gemma Salvadó, Shorena Janelidze, Sebastian Palmqvist, Niklas Mattsson-Carlgren, Piero Parchi, Henrik Zetterberg, Nicholas J Asthon, Oskar Hansson

## Abstract

Older individuals frequently harbor multiple brain pathologies, including Alzheimer’s disease (AD) related amyloid-β (Aβ) and tau alongside α-synucleinopathy and vascular pathology. Proteomic profiling offers a strategy to better understand common as well as unique features of these different brain pathologies.

We analyzed cerebrospinal fluid (CSF) (n=1,658) and plasma (n=749) samples from participants in the BioFINDER cohorts using the automated NULISAseq CNS Disease panel of 125 proteins. Differentially abundant proteins (DAPs) related to AD pathology (based on Aβ– and tau-PET positivity), α-synuclein (based on synuclein amplification assay [SAA] positivity) and vascular pathology (based on white matter lesion [WML] load) were identified with linear models simultaneously including a binary measure for the three pathologies. In the BioFINDER-2 subcohorts, DAPs were further evaluated for associations with continuous baseline (n=1,137) and longitudinal (n=656) Aβ-PET, tau-PET, and WML measures in models accounting for all pathologies. Associations with AD-signature cortical atrophy (n=915) and cognitive decline by the MMSE (n=1054) were also examined.

We identified 84 CSF DAPs, with largely distinct protein signatures for each pathology (AD, n=66 DAPs; vascular pathology, n=55; α-synuclein pathology, n=16). 10 DAPs (e.g., FABP3, UCHL1, NPTXR, NPTX2) were altered across all three pathologies, reflecting general neurodegeneration. AD-associated DAPs included glial/inflammatory markers (CHIT1, CX3CL1, CD63) linked to Aβ pathology, and synaptic/neuronal injury markers (VSNL1, NRGN, NEFL) and metabolic enzymes (FABP3, MDH1) linked to tau pathology. Aβ-associated proteomic differences were most evident in CU individuals, while tau-associated differences predominated in MCI. More proteins, particularly neurodegeneration and synaptic markers, were associated with tau change than with Aβ change. Vascular pathology exhibited a distinct profile, enriched for inflammatory, angiogenic and extracellular matrix proteins (PGF, POSTN, TREM1, VCAM1). DDC was the main protein associated with α-synucleinopathy. Only a few proteins, including UCHL1, NPTX2, and NEFL, predicted cognitive decline and cortical atrophy after accounting for all brain pathologies.

In plasma, although fewer DAPs were identified (n=20), findings included established AD biomarkers. Only plasma VCAM1 and NEFL were associated with α-synuclein and vascular pathology.

NULISA identified stage-dependent, disease-specific CSF biomarker signatures with limited overlap, alongside shared neurodegenerative markers, supporting improved biological interpretation and more refined classification of neurodegenerative pathology.

## Introduction

Data from community-based neuropathological studies indicate that multiple brain pathologies are highly prevalent in older adults, often even before cognitive symptoms become apparent (1–3). Common neurodegenerative proteinopathies include amyloid-β (Aβ) plaques, tau neurofibrillary tangles, and α-synuclein inclusions, with reported frequencies of Alzheimer’s disease (AD) related pathology of approximately 19–67% and Lewy body pathology of 6–39% at autopsy in older individuals irrespective of the clinical symptoms (1). Cerebrovascular lesions, including micro-and macroinfarcts and white matter lesions (WML), are also frequent, with vascular pathology present in roughly 28–70% of cases (2, 3). Mixed pathology, defined as the co-occurrence of multiple pathologies, is more common than a single, isolated pathology (4, 5) and appears to exert additive or synergistic effects on neurodegeneration and clinical presentation (6), complicating both diagnostic interpretation and the development of targeted treatments.

AD pathology characterized by the presence of Aβ plaques and tau neurofibrillary tangles (7). The development of *in vivo* fluid biomarkers for these proteins has greatly benefited the diagnosis and staging in research settings (8, 9). In contrast, in patients with suspected neurodegenerative diseases, recommended approaches for assessing in vivo non-AD pathologies are limited to the α-synuclein seed amplification assay (αSyn-SAA) and the detection of infarcts and WML by magnetic resonance imaging (MRI). These approaches increase clinical cost and burden. A fluid biomarker-based multi-analyte assay covering all major categories and pathologies would represent a far more efficient and comprehensive strategy (7, 10). However, large-scale studies evaluating fluid biomarkers for vascular lesions and α-synuclein pathology remain scarce, leaving important gaps in both diagnostic approaches and understanding of the underlying biological processes across multiple pathologies.

While studies from our lab and others have explored distinct proteomic profiles across the AD continuum (11–14) and various manifestations of cerebral small vessel disease (cSVD) (15) in CSF, these analyses have primarily relied on large scale proteomics platforms such as Olink and SomaScan. Although these platforms enable proteomic profiling to uncover molecular and pathological mechanisms, they may introduce variability due to suboptimal reagent performance in multiplexed settings (16). Alternatively, the newly developed Nucleic Acid Linked Immuno-Sandwich Assay (NULISA™) offers attomolar sensitivity in a mid-scale proteomics platform (17). The proteins included in its panels are selected for their relevance to neurodegenerative diseases and inflammation including those with post-translational modifications that are not always well covered by other methods. However, their relationships to other pathologies beyond AD are not yet fully established.

Altogether, this study aims to identify CSF and plasma proteins within the NULISA platform that can differentiate Aβ, tau, α-synuclein, and vascular pathology. By evaluating these proteins, we aim to characterize their relationships to each pathology, explore co-expression patterns, and gain insight into the underlying biological processes.

## Methods

### – Participants

We included participants from the ongoing prospective Swedish BioFINDER-1 (n=47; NCT01208675) and BioFINDER-2 cohorts (n=1611; NCT03174938). The participants ranged from adults with intact cognition or subjective cognitive decline (SCD) (both included in the cognitively unimpaired [CU] group), mild cognitive impairment (MCI), and dementia. Patients referred to the memory clinic with cognitive symptoms were classified as having MCI if they scored ≥1.5 SD below age– and education-adjusted norms in any cognitive domain from an extensive neuropsychological battery, including verbal fluency, episodic memory, visuospatial ability, and attention/executive domains (described in detail for BioFINDER-1(18) and BioFINDER-2(6)). Those not meeting MCI criteria were classified as having SCD. AD participants were required to be Aβ-positive and to meet the DSM-5 criteria for dementia due to AD (19, 20). The non-AD group included patients with Parkinson’s disease (PD), Lewy body dementia (LBD), and vascular dementia (VaD). All participants have been recruited from the general population, at the Memory Clinic or Neurology clinic at Skåne University Hospital or Ängelholm Hospital in Southern Sweden from 2008 to 2023. The main inclusion and exclusion criteria, as well as the clinical diagnostic criteria for each disease, have been described in detail previously (21) and on https://biofinder.se. All participants included in the study had all the following data available at baseline: tau-PET, structural MRI, CSF α-Syn Real-Time Quaking-Induced Conversion (RT-QuIC) and CSF Aβ42/Aβ40 or Aβ-PET data.

### – Cognition

In BioFINDER-2, cognitive decline was assessed using both the Mini-Mental State Examination (MMSE) and the modified Preclinical Alzheimer’s Cognitive Composite (mPACC5). The mPACC5, designed to capture early cognitive changes, was analogous to the original PACC5(22). It was computed as the average of four z-scores: memory performance (delayed recall from the Alzheimer’s Disease Assessment Scale-Cognitive Subscale [ADAS-Cog], double-weighted to maintain the emphasis on memory in the original PACC), verbal fluency (animal fluency), executive function (Symbol Digit Modalities Test), and global cognition (MMSE), as previously described (23).

### – Magnetic resonance imaging

Participants were examined using a Siemens 3 T Trio scanner (Siemens Medical Solutions) in BioFINDER-1 or a Siemens 3 T MAGNETOM Prisma scanner (Siemens Medical Solutions) in BioFINDER-2, as described previously (21, 24).

Imaging included a whole-brain T1-weighted anatomical magnetization-prepared rapid gradient echo sequence (MPRAGE; TR = 1900 ms, TE = 2.54 ms, voxel size = 1 × 1 × 1 mm3) and a T2-weighted fluid-attenuated inversion recovery sequence (FLAIR; TR = 5000 ms, TE = 393 ms, matching resolution with the T1-weighted image). Cortical reconstruction and volumetric segmentation were performed using the FreeSurfer image analysis pipelines (v6.0). White matter lesions (WML) were segmented and quantified using the longitudinal Sequence Adaptive Multimodal SEGmentation (SAMSEG) tool from FreeSurfer (version 7.1), with T1-weighted and FLAIR images as inputs (25, 26). WML volume was adjusted by individual intracranial volume (ICV). Whole-brain volume was calculated as the sum of grey and white matter volumes minus ventricular volume, also adjusted for ICV. Cortical thickness was defined as the distance between the gray/white matter boundary and the pial surface. The AD-signature cortical thickness measure included the bilateral entorhinal, inferior temporal, middle temporal, and fusiform cortices (27).

### – PET acquisition and processing

Aβ– and tau-PET scans were acquired using a GE Discovery MI digital scanner (General Electric Medical Systems). Tau-PET was conducted 70-90 minutes after injecting ∼370 MBq of [^18^F]RO948, while Aβ-PET was performed 90-110 minutes post-injection of ∼185 MBq of [^18^F]flutemetamol (21). Standardized uptake value ratios (SUVRs) were calculated using the inferior cerebellum as the reference region for tau-PET and the whole cerebellum for Aβ-PET. Aβ-PET SUVR was computed for a neocortical composite region encompassing the prefrontal, lateral temporal, parietal, anterior cingulate, and posterior cingulate/precuneus regions. Previously reported positivity cutoffs, established using Gaussian mixture modeling (28), were ≥1.033 SUVR for BioFINDER-2 and ≥1.138 SUVR for BioFINDER-1. For tau-PET, a temporal lobe composite SUVR was derived from the entorhinal cortex, inferior/middle temporal, fusiform gyrus, parahippocampal cortex and amygdala. This composite reflects Braak stages I-IV. Participants were considered tau-positive if ≥1.362 SUVR, based on Gaussian mixture modeling as previously reported (28).

### – Blood and CSF sample collection and processing

20 mL of CSF was collected in 5 mL LoBind tubes. CSF was centrifuged (2000g, +4 °C) for 10 min, aliquoted in 1.5 mL polypropylene tubes and stored at −80 °C within 30-60 min of collection (29). Blood was collected in EDTA-plasma tubes (Vacutainer K2EDTA tube, BD Diagnostics) and centrifuged (2000g, +4 °C) for 10 min. Resulting plasma was transferred into one 50 mL polypropylene tube, mixed and aliquoted into 1.5 mL polypropylene tubes and stored at −80 °C within 30-60 min of collection. CSF and plasma samples were collected during baseline visits for BioFINDER-2 participants, while for BioFINDER-1 participants, samples were obtained at follow up visits. CSF Aβ42/40 ratio was predominantly quantified using the Elecsys immunoassay, with a predefined cutoff of 0.08 to define Aβ-positivity (21). Few cases did not have Elecsys measurement available in which routine clinical Lumipulse G assay results were used instead, applying a predefined cutoff of 0.072 (30) (30). The presence of α-synuclein aggregates was assessed using the αSyn-SAA performed on CSF samples, implemented according to a previously established RT-QuIC protocol (31). Measurements were done blinded to diagnostic categories.

### **-** Pathological classification criteria

For each participant, we derived a binary indicator of the presence of AD, α-synuclein and vascular pathology, independent of clinical diagnosis.

AD pathology was defined by the presence of both Aβ and tau positivity. Aβ positivity was primarily determined using Aβ-PET. By study design Aβ-PET is not done for patients AD dementia or other neurodegenerative diseases (n=460), and thus Aβ positivity was instead determined by the CSF Aβ42/Aβ40 ratio for such participants. Tau positivity was determined using tau-PET, using a priori predefined cutoff previously published (28).

Vascular pathology was defined based on WML load. Participants were classified as having vascular pathology if their WML load fell within the highest tertile, with a cutoff of 0.594% of ICV. This data-driven threshold is cohort-dependent and relatively conservative, reflecting a high lesion burden and therefore identifying individuals with substantial vascular involvement and has been previously described (15, 32, 33).

Participants were classified as having α-synuclein pathology if the CSF αSyn-SAA test was positive in 3 or 4 of 4 replicates, or in > 33% of replicates (i.e., in ≥4 of 12) when the sample required retesting (for details see (34)).

### **-** NULISA proteomics measures

Samples were randomized across plates, and quality control (QC) samples were included throughout. CSF samples from all individuals were analyzed using the NULISAseq CNS Disease Panel (Almar Biosciences), which measures 125 unique proteins across seven categories: Aβ and tau pathology, inflammation, neurodegeneration, synuclein and synaptic disorders, vascular pathology, and metabolism. In a subset of 749 individuals, CSF was also analyzed using the NULISAseq Inflammation Panel (Alamar Biosciences), expanding the total number of unique proteins analyzed to 324. Additionally, in the same 749 individuals, plasma was analyzed using the NULISAseq CNS Disease Panel. For an overview of the participants, samples and biomarker panels used in each analysis see Supplemental Figure 1. Protein concentrations were reported as NULISA Protein Quantification (NPQ) units, derived from normalizing sequence quantification counts for intraplate and intensity variability, followed by log2 transformation to approximate normality. Analyses were conducted as previously described (35).

Each protein target had a plate-specific limit of detection (LOD), determined by adding three times the standard deviation (SD) of the unlogged counts from negative controls included in each plate. To ensure robust analysis, we restricted our dataset to proteins with widespread expression, defined as those detected above the LOD in at least 70% of participants. Only data points which failed quality control were excluded from analysis, while data below LOD were retained in analysis. This resulted in the retention of 106 proteins in CSF and 116 in plasma in the CNS panel for the main analysis. In the 749 individuals with both CNS and Inflammation panel, 236 proteins were retained for subsequent analysis, 102 proteins from the CNS panel and 132 from the Inflammation panel. Additionally, based on previous findings from our lab highlighting the impact of inter-individual variability on CSF protein concentrations, largely influenced by CSF dynamics and differences in clearance and production rates (36), we calculated a measure of mean overall protein levels to use as a covariate in our analysis. This measure was derived by averaging the z-scored NPQ values of highly detected proteins, defined as those exceeding the LOD in over 90% of participants (95 in CSF for the CNS panel, 114 proteins in plasma for the CNS panel and 202 in CSF for the subanalysis including both the CNS and Inflammation panels). Measurements were done blinded to diagnostic categories. Additional File 1 contains separate worksheets for each analysis, listing all proteins included and their corresponding associations.

### – Statistical analysis

All analyses were performed in R v.4.4.2 or Python v.3.12.6. All plots were generated with the R package ggplot2 v.3.5.1. All statistical tests were two sided and P values adjusted for FDR.

#### – Differential abundant protein analysis

For each protein, we applied a linear regression model with protein abundance as outcome, to identify differentially abundant proteins (DAPs) across pathologies. This model simultaneously included binary measures (presence vs. absence) of AD, α-synuclein, and vascular pathology as predictor variables while adjusting for age, sex, and overall protein levels as covariates. Predictors, outcomes and covariates were all z-scored to facilitate comparability across measures. Multiple comparisons were controlled for using the Benjamini-Hochberg method (FDR), and we focused on DAPs with P_FDR_<0.05 for subsequent analyses.

As a sensitivity analysis, we also ran separate linear regression models for each pathology independently, each adjusted for age, sex, and overall protein level. We compared the sets of DAPs identified in these independent models to those from the combined model to evaluate the robustness of our findings and confounding effects of co-pathologies.

#### – Associations with continuous measures of pathology

Next, we examined associations between the identified DAPs and continuous measures of Aβ-PET, tau-PET, and WML load both at baseline and their rate of change over time. The cross-sectional analysis was performed in the BioFINDER-2 CU and MCI participants (n = 1173) where all participants had baseline Aβ-PET and tau-PET data. Analyses were additionally conducted stratified by cognitive status. Participants with dementia were excluded, as they did not undergo Aβ-PET by study design. Out of those, 656 also had longitudinal data available for three of the pathological features (Aβ, tau and WML). For these participants, individual rates of change in continuous measures were estimated using linear mixed-effects models with random intercepts and slopes, implemented in the *lme4* package (v1.1-36) in R. PET SUVR values or WML volumes were modeled as the dependent variable, with time (years since baseline) as the only predictor. Participant-specific slopes, representing annual change in Aβ– and tau-PET SUVR and WML volume, were extracted from these models. Each participant contributed between two and six follow-up scans for Aβ– and tau-PET (median [IQR]: 1.97 [2.09] and 2.00 [2.01] years, respectively) and between two to six MRI scans (median [IQR]: 2.04 [2.02] years).

Linear regression models were used to investigate the associations between either baseline levels of pathology or rates of change in pathology (all pathologies included as predictors) and protein abundance as the outcome. Age, sex, and overall protein levels were also included as predictors, as well as α-synuclein positivity for the cross-sectional associations. Similarly, analyses were conducted in the whole subset of individuals and stratified by cognitive status.

#### – Inferred trajectory using pseudotime

We constructed a pseudotime variable to model AD progression as a continuous measure based on pathology to visualize protein trajectory along this pseudotime. Briefly, we derived the AD pseudotime using the full BioFINDER-2 sample (n=1504) with the necessary input data, as described previously (11). Plasma tau phosphorylated at Thr217, a biomarker strongly correlated with brain Aβ pathology load, was combined with tau-PET SUVR values from three meta-ROIs corresponding to different Braak stages to model fibrillary Aβ and tau pathology. In short, each participant was assigned a pseudotime score ranging from 0 (minimal pathology) to 1 (highest pathology) using trajectory inference method in Phyton. To visualize protein trajectories in relation to disease progression, we applied a smoothed generalized additive model (GAM) in ggplot2, allowing for flexible trend estimation across the AD progression. WML associations were also modeled with GAM, using WML load as the independent variable.

#### – Cell-type enrichment analysis

We assessed the differential expression of genes of interest across cell types using the same approach as a recently published study (11). Specifically, we utilized RNA sequencing (RNA-seq) data from the middle temporal gyrus (MTG) provided by the Allen Brain Atlas consortium. The Seurat object (MTG10xSEA-AD2022, https://portal.brain-map.org/atlases-and-data/rnaseq/human-mtg-10x_sea-ad) containing single-nucleus transcriptomes from over 1.6 million nuclei across five postmortem human brains was downloaded (37). To quantify cell-type-specific expression, we applied the AverageExpression function from the R package Seurat v.4.3.0, which calculates mean expression levels using the class and subclass annotations available in the Allen Brain dataset (38). This analysis included both neuronal (GABAergic and glutamatergic neurons) and non-neuronal cell types (microglia, astrocytes, oligodendrocytes, VLMC, endothelial and OPCs), while excluding nuclei labeled as “None.” Finally, we computed each cell type’s contribution as its expression proportion relative to the total expression across all cell types.

#### – Functional enrichment analysis

We used the WEB-based Gene SeTAnaLysis Toolkit (WebGestalt) for functional enrichment analysis on identified DAPs (39). We performed human over-representation analysis using gene ontology (GO) database for biological process (BP), defining our background set to the whole protein coding genome. To reduce term redundancy, we used the affinity propagation option. We performed functional enrichment analysis separately for upregulated and downregulated DAPs. However, due to the low number of upregulated proteins linked to α-synuclein pathology, which was not sufficient for a separate analysis, all α-synuclein-related proteins were analyzed together as a single group. P-values were adjusted using Benjamini-Hochberg method.

#### – Cognitive decline and cortical atrophy prediction

For longitudinal prediction analyses, participant-specific annualized slopes for AD-signature cortical thickness, whole-brain volume, and cognitive scores (MMSE and mPACC) were estimated using linear mixed-effects models with random intercepts and slopes, as described above. These individualized slopes were then used as dependent variables in separate linear regression models for each protein. All models were adjusted for age, sex, and mean CSF protein concentration, with years of education additionally included in the cognitive models. Extended models further incorporated baseline pathology measures (Aβ– and tau-PET SUVR, WML volume, α-synuclein positivity) to assess the independent contribution of protein levels to change in cognition and brain structures over time. Analyses were additionally conducted stratified by cognitive status.

### – Code availability

All code was written using available packages in R and Python and can be provided by contacting the authors.

## Results

### 1. Participants

We included 1658 participants from the Swedish BioFINDER-1 and 2 cohorts, representing individuals across the AD continuum as well as other neurodegenerative conditions, including VaD, PD and LBD.

Participants were classified based on the presence of each possible measured pathology, with multiple pathologies possible per individual, as described in the Methods. Among the 1658 participants, 368 had AD pathology, 314 had α-synuclein pathology, and 546 had vascular pathology (Table 1). 763 individuals showed no evidence of any pathology, whereas 40 had all three pathologies. From the combination of two pathologies, having both vascular and AD pathology was the most frequent (n=121 participants), followed by vascular and α-synuclein pathology (n=89), and α-synuclein and AD (n=43). For details on participants with each pathology refer to Supplemental Table 1.

**Table 1:**
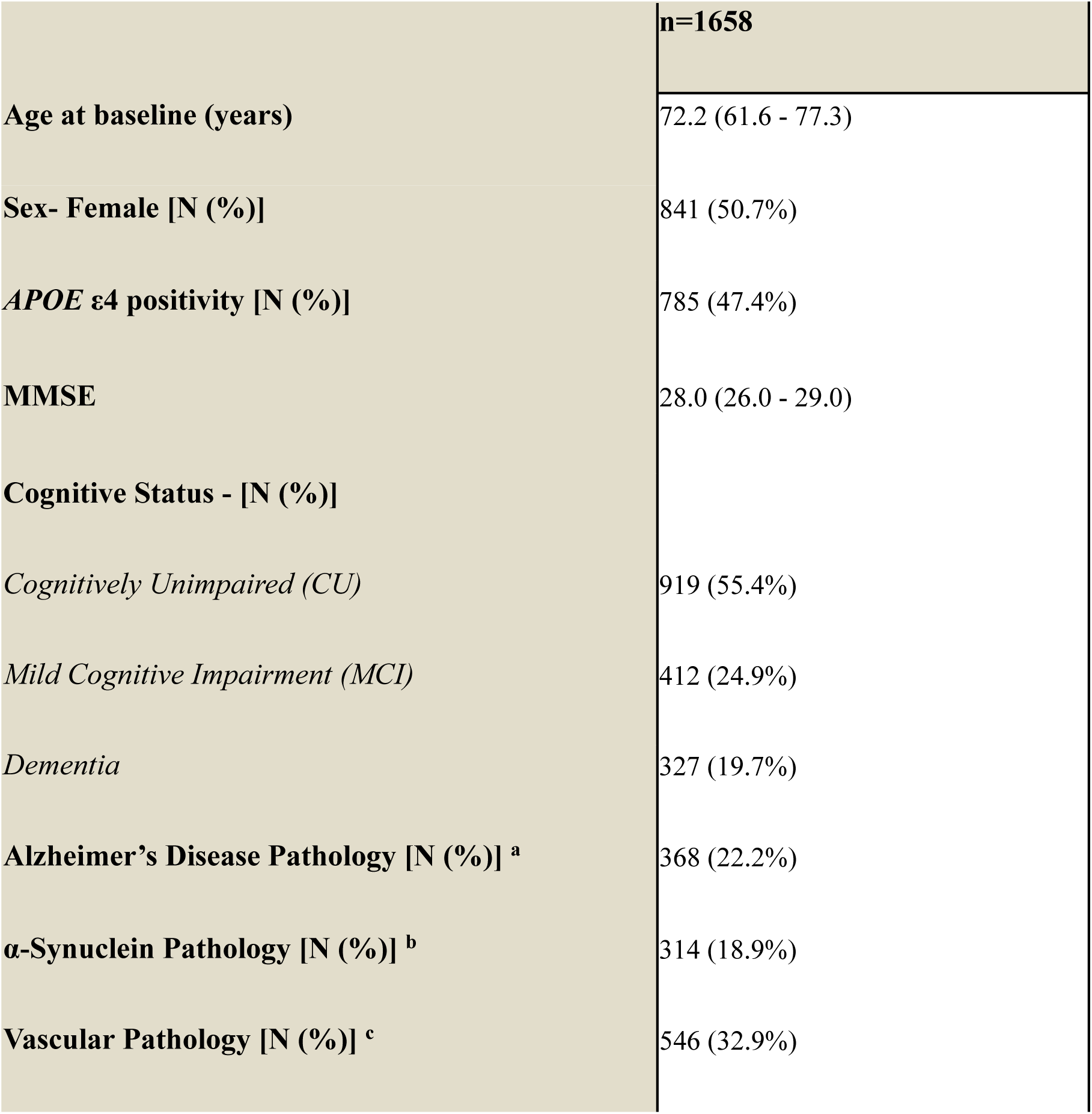
Demographics of included participants. Data shown as mean (95%CI) unless specified otherwise. Abbreviations: APOE, apolipoprotein E; CU, Cognitively Unimpaired; MCI, Mild cognitive impairment; MMSE, Mini-Mental State Examination; SUVR, standardized uptake value ratio. a. Aβ positivity determined by [^18^F]flutemetamol SUVR >1.033 (BioFINDER-2) or >1.138 (BioFINDER-1), or CSF Ab42/40 < 0.072 (Lumipulse G). Tau positivity defined by [^18^F]RO948 tau-PET Braak I-IV SUVR > 1.362 b. Synuclein pathology positivity determined by positive α-synuclein seed amplification assays (SAA). c. WML positivity set at >0.59% of intracranial volume (ICV), top tertile

### 2. Shared biomarker profiles

We first identified DAPs associated with each pathology while adjusting for the presence of the other two pathologies. In total, we identified 84 DAPs in CSF across the three pathologies, with most proteins related to vascular and AD pathology (Fig. 1a). Among the 84 identified DAPs, only 10 were shared across all three pathologies (Fig. 1b): FABP3 and UCHL1 were increased whereas AGRN, Aβ38, Aβ40, Aβ42, NPTX2, NPTXR, TAFA5 and VEGFA were decreased in all pathologies. While many proteins were unique to a single pathology, 19 DAPs were shared between two pathologies, showing same directional changes (i.e., increased or decreased abundance in both; Fig. 1b): in both AD and α-synuclein pathology CX3CL1, IFNG, NPY and PSEN1 were decreased. In AD and vascular CCL3, CHIT1, GFAP, NEFL and TREM1 were increased, while BACE1, IGF1R, IL9, NPTX1 and TEK1 were decreased. In α-synuclein and vascular pathology, IL7 and NEFH were increased, while CRH, GOT1, and SOD1 were decreased. Additionally, we identified 22 DAPs with inverse associations between pathologies (Fig. 1c), most of which were increased in AD but showed decreased abundance in vascular pathology: ACHE, APOE4, MAPT, MDH1, NRGN, SLIT2, SNCB and p-tau forms.

**Figure 1:**
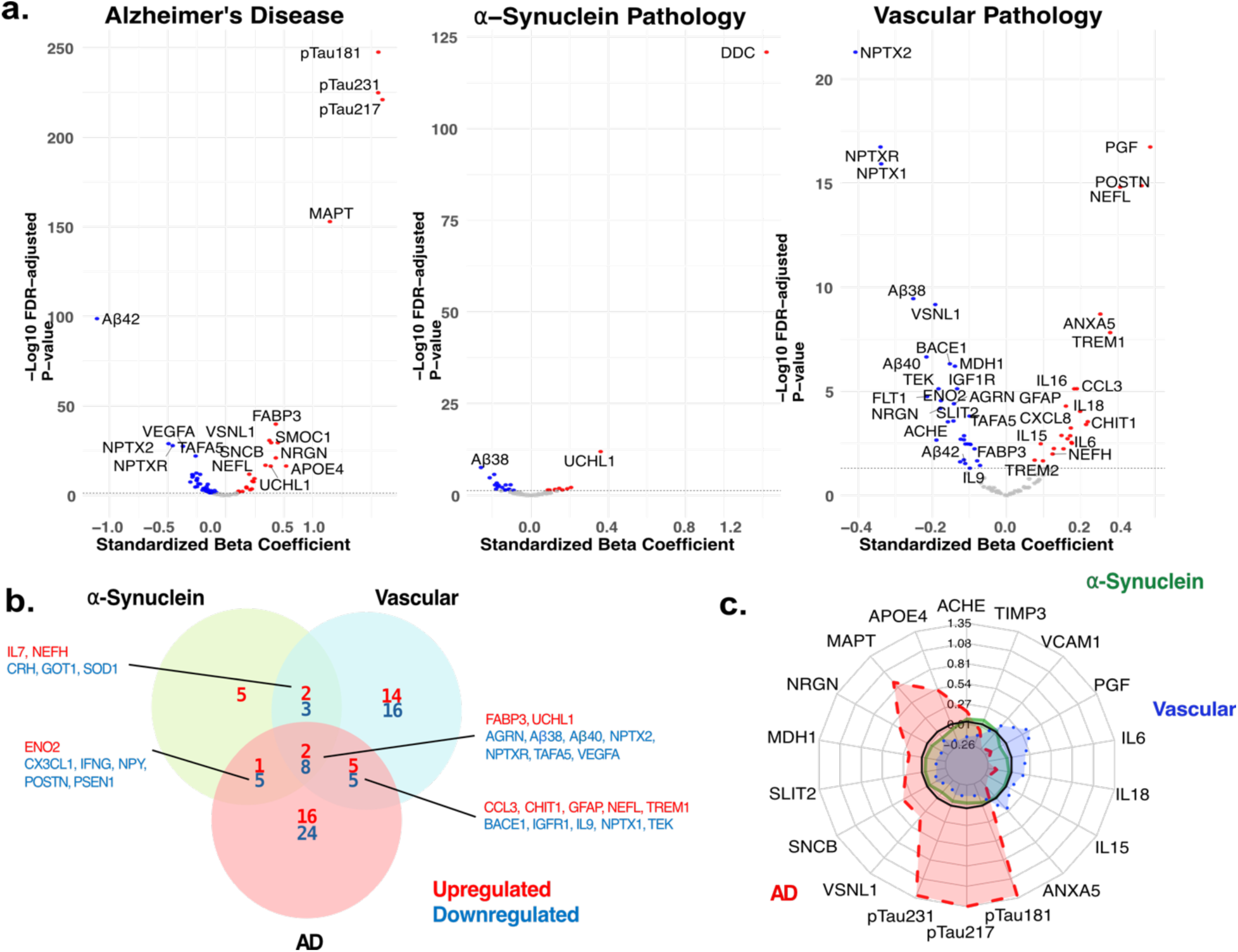
Identification of Differential Abundant Proteins in CSF across different diseases. **a**) Volcano plots show proteins linked to AD, α-synuclein, and vascular pathology in CSF, analyzed together. The x-axis represents standardized β values (effect size), and the y-axis shows –log10 p-values (statistical significance). Model adjusted for age, sex, and CSF protein levels. Dashed lines mark significance at α=0.05 (FDR-corrected). Significant proteins (p[FDR]<0.05) are in red (higher abundance in presence of pathology) or blue (lower abundance). **b**) Venn diagram shows shared and unique proteins linked to AD, ⍺-synuclein, and vascular pathologies, with red and blue indicating positive and negative associations, respectively. c) Radar chart displays standardized β coefficients of shared proteins across pathologies, with each axis representing a protein. The black circular axis marks βstd=0

P-tau217, p-tau181, and p-tau231 were the proteins with the strongest association with AD pathology, showing increased abundance (1.31 < std β < 1.35, p < 0.001; Fig. 1a). Although a greater number of proteins were associated with vascular pathology, their standardized beta coefficients were generally smaller than those for AD and α-synuclein pathology. NPTX2, NPTX1, and NPTXR were less abundant with vascular pathology (−0.41 < std β < –0.34, p < 0.001), whereas PGF, NEFL, and POSTN showed increased abundance in the presence of vascular pathology (0.31 < std β < 0.39, p < 0.001). DDC showed the strongest and most distinct association with α-synuclein status (std β= 1.22, p < 0.001). FGF2, the second DAP with the strongest specific association to α-synuclein, was considerably lower (std β= 0.36, p < 0.001).

In a smaller subset of the sample (n=749), the same CNS panel was available in plasma samples (Supplementary Table 2). Repeating the same analyses, we found 20 DAPs in plasma out of the 84 DAPs identified in CSF, with most associated with AD pathology (Supplemental Fig. 2a). Inflammation and CNS panels were available for CSF, only in this same subset, including a total of 236 proteins. Here we identified 84 DAPs, of which 48 overlapped with the main analysis. From the inflammation panel, most of the DAPs were associated with vascular pathology, including proteins such as MMP12, TIMP1, CD4 (0.24 < std β < 0.44, p < 0.001), TNFRSF18 and CD46 (std β= –0.28 and –0.21, p<0.006) (Supplemental Fig. 2b). The strongest associations largely remained within the CNS panel.

### 3. Cell-type and functional enrichment of protein signatures

Out of the ten proteins commonly dysregulated across pathologies, nine (ACHE, AGRN, ENO2, FABP3, NPTX2, NPTXR, POSTN, TAFA5, UCHL1) were mainly expressed in neurons (Fig. 2a). These proteins likely reflect downstream neuronal dysfunction independent of the underlying cause. Consistently, functional enrichment analysis identified terms related to neuronal injury and synaptic dysfunction across all pathologies (Fig. 2b). VEGFA, also commonly altered, was enriched in astrocytes and oligodendrocyte precursor cells (OPCs), suggesting involvement of glial-vascular pathways.

**Figure 2:**
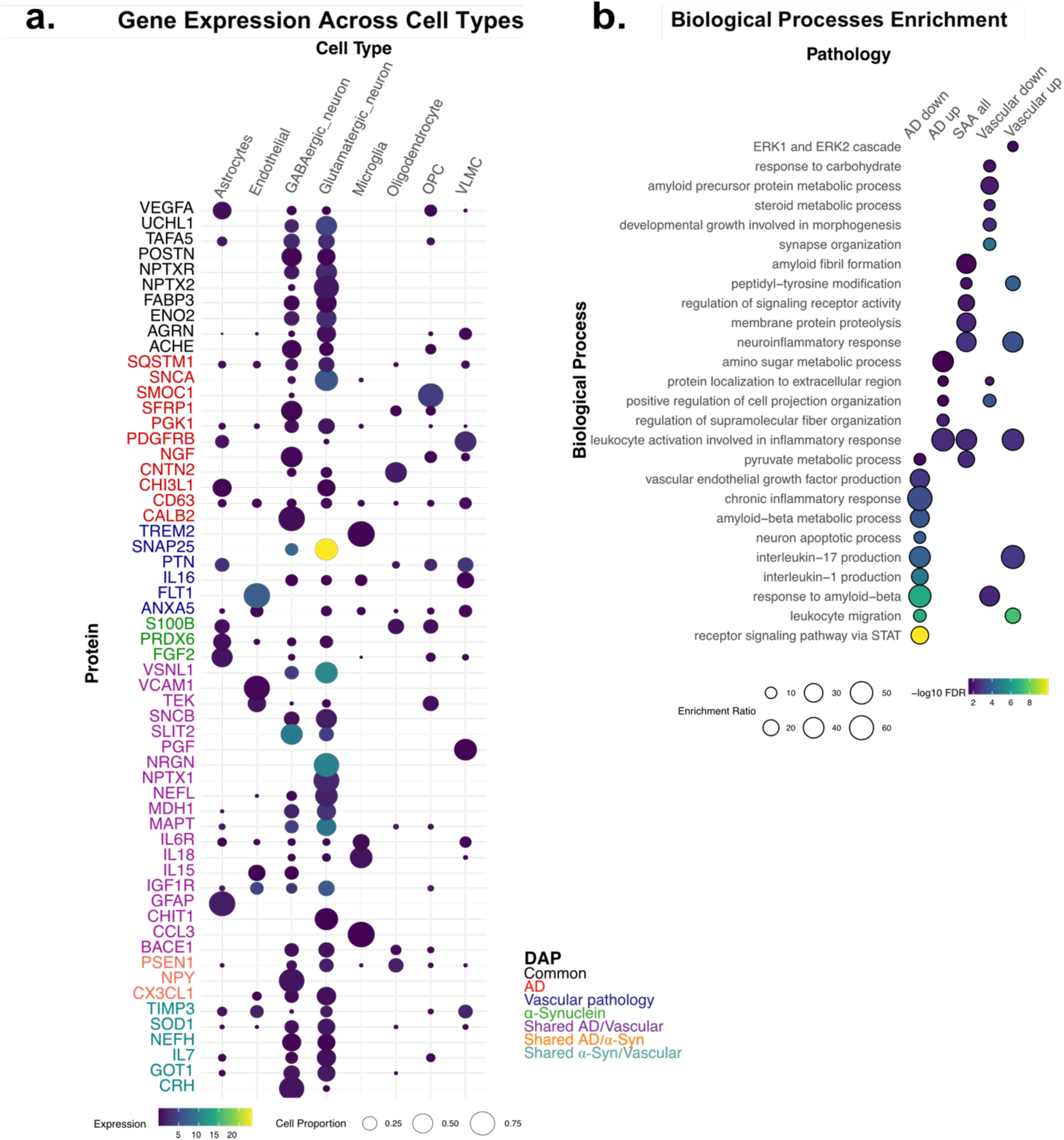
Cell-type and biological process enrichment in differential abundant proteins. **a)** Cell-type-specific expression of differentially abundant protein transcripts. Only transcripts previously identified as DAPs with an average expression >0.05 and detected in at least 5% of cells in any of the analyzed cell types are shown. Bubble size reflects the proportion of cells within each type expressing the transcript; color indicates average expression level. Protein names on the x-axis are colored according to the pathology with which the protein was identified as a DAP. Data are from single-cell transcriptomics of the human middle temporal gyrus (MTG). **b)** Significant Gene Ontology (GO) Biological Process terms enriched in DAPs for each pathology, using the protein-coding genome as background for enrichment analysis. Bubble size reflects the enrichment ratio of biological process; color corresponds to the significance (p[FDR]) of the enrichment.

Proteins associated with AD pathology were predominantly expressed in neurons, with notable exceptions such as GFAP and CHI3L1 (YKL-40), which were mainly expressed in astrocytes, SMOC1 predominantly in OPCs, CNTN2 in oligodendrocytes and PDGFRB in astrocytes and vascular and leptomeningeal cells (VLMC). Correspondingly, functional enrichment revealed Aβ-related processes, cytoskeletal remodeling, and extracellular protein localization, alongside immune and inflammatory signaling, suggesting concurrent synaptic disruption, aggregate deposition, and glial activation. Enrichment of amino sugar and pyruvate metabolism further supports neuronal stress and impaired energy metabolism in AD (Fig. 2b).

Vascular pathology showed strong enrichment of proteins expressed in endothelial cells (FLT1, VCAM1, VSNL1), microglia (TREM2, interleukins), VLMCs (PGF, PTN, TIMP3), and OPCs (PTN, TEK), indicating involvement of vascular, immune, and supporting glial compartments (Fig 2a). Pathway analyses revealed significant enrichment for extracellular matrix remodeling, angiogenesis, and synaptic organization. Metabolic terms related to lipid handling also appeared, pointing to metabolic stress potentially increasing neuronal vulnerability (Fig. 2b). Together, these findings support a model in which vascular injury triggers coordinated inflammatory, structural, and metabolic responses across multiple cell types.

DAPs associated with α-synuclein pathology were primarily enriched in astrocytes (Fig. 2a). The functional enrichment profile was dominated by immune, inflammatory, signaling, and metabolic processes consistent with astrocyte-driven neuroinflammatory responses (Fig. 2b).

### 4. Associations with the severity of pathology load

Next, we assessed whether the identified CSF DAPs varied in response to the load of Aβ plaque pathology (Aβ-PET SUVR), tau tangle pathology (tau-PET SUVR), and WML load (MRI WML volume), and the presence of α-synuclein pathology. We included all pathologies in a single linear model across all BioFINDER-2 participants for which all these data were available (n = 1173, Supplemental Table 3), as well as separately in CU and MCI groups to determine whether the protein associations with pathology differed by clinical stage. An overview of protein associations for each pathology, showing color-coded directionality and key hits, is presented in Supplemental Figure 3.

In the main analysis, FABP3 and UCHL1 emerged as DAPs associated with all pathologies (Fig. 3a). However, stratified analysis in CU and MCI groups revealed that no DAPs were common across all four pathology types (Supplemental Fig. 4). Overlapping proteins were found between Aβ– and tau-PET and WML load: p-tau species, MAPT, NRGN, PTN and AGRN were associated with all 3 continuous pathologies in the whole population, but only p-taus species and MAPT survived the stratified analysis in both CU and MCI. The largest intersection was between Aβ-PET and WML load. The overlapping proteins related to both Aβ-PET and WML included: apoE, SLIT2, ANXA5, CXCL8 and PDGFRB, and related to both tau-PET and WML included VSNL1, ENO2, MDH1, ACHE, CCL17, PGF, IL16, IL15, IL6, IL18, CSF2, with some of them going in opposite directions. Relatively few proteins were negatively associated with all pathological loads, AGRN consistently showed negative associations with Aβ-PET, tau-PET, and WML, while others such as PTN, TEK, VEGFA, TAFA5, IL5, BACE1, Aβ40, Aβ38, and neuropentraxins were negative only in Aβ-PET and WML.

**Figure 3.**
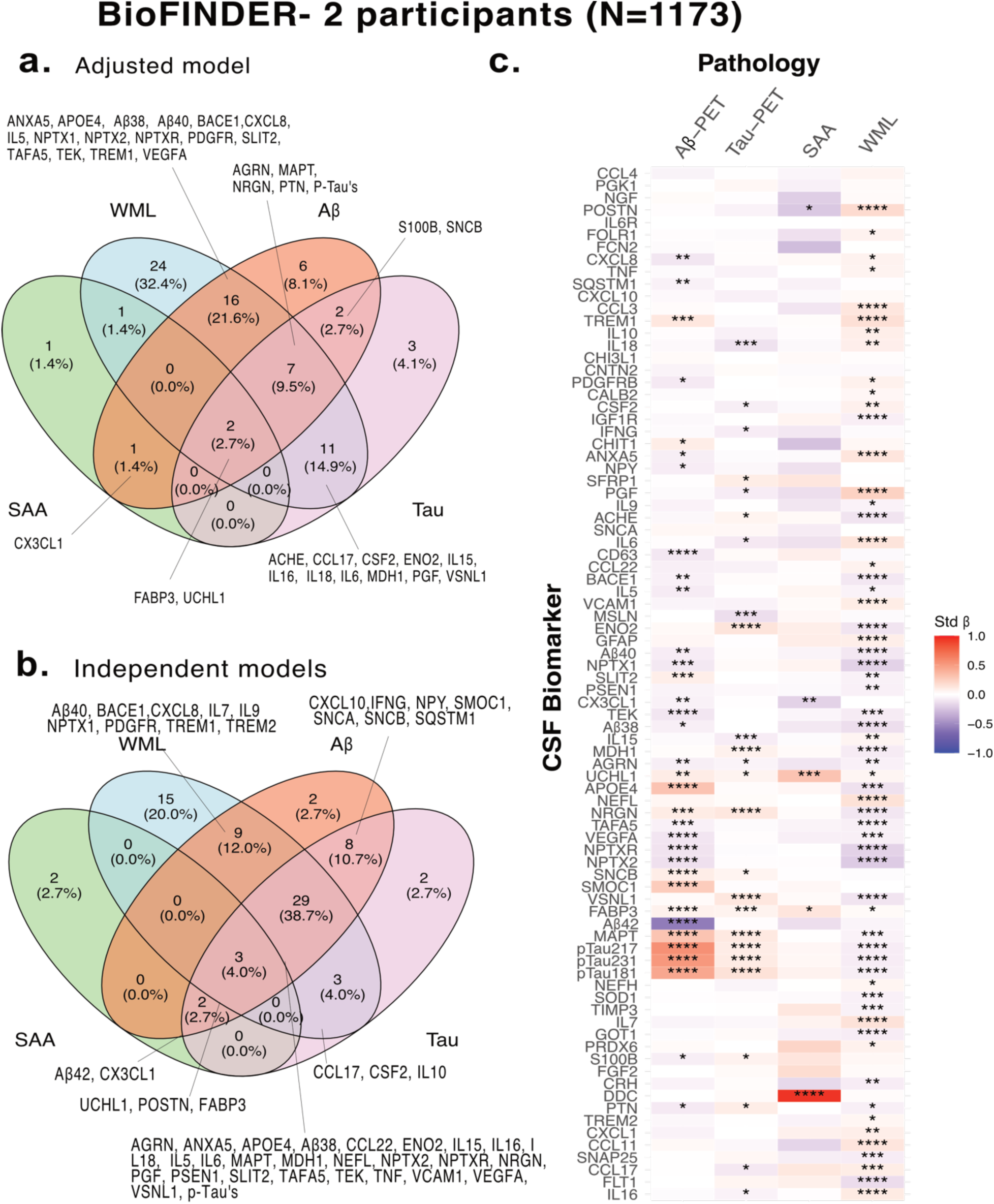
Associations between DAPs and the load of pathology and presence of ⍺-synuclein pathology Panels. **a-b** show Venn diagrams of DAPs associated with burden of Aβ, tau, ⍺-synuclein and WML pathology. **(a)** display results from a single mutually adjusted model including all four pathologies whereas **(b)** show results from independent models for each pathology. **Panel c** presents a heatmap illustrating standardized beta coefficients from the mutually adjusted model for all four pathologies for all participants. Cell color reflects the direction (red for positive, blue for negative association) and strength (intensity of color proportional to absolute beta value) of each association between protein concentration and pathology burden. All models were adjusted for age, sex, and CSF protein levels. Analysis focused on previously identified DAPs. \**P_FDR_* < 0.05, *\*\*P_FDR_* < 0.01, *\*\*\*P_FDR_* < 0.001, *\*\*\*\*P_FDR_* < 0.0001.

Sensitivity analyses revealed that more proteins reached significance when models were run independently by pathology (Fig. 3b); mutual adjustment reduced these associations, highlighting confounding effects (Supplemental Fig. 5). These effects were most pronounced for tau-PET associations. Alternatively, some protein associations became significant only once adjusting for other pathologies, like PTN and S100B for Aβ– and tau-PET associations.

Distinct marker profiles emerged for each pathology. Several proteins were exclusively associated with Aβ-PET load such as SMOC1, CHIT (0.09 < std β < 0.30, p < 0.03), Aβ42, CD63, SQSTM1 and NPY (−0.57 < std β < –0.06, p < 0.02; Fig. 3c), which had in general larger associations than in CU participants (Supplemental Fig. 4). For tau-PET load, the following proteins were uniquely changed: MSLN, IFNG (std β= –0.13 and –0.08, p<0.05, respectively) and SFRP1 (std β = 0.10, p < 0.02; Fig. 3c). Additionally, proteins including ENO2, VSNL1, NRGN, and IL15 were consistently associated with tau-PET load across both CU and MCI groups, whereas MDH1 showed a unique association in the MCI group.

Notably, Aβ-PET-associated DAPs predominated in CU, while tau-PET DAPs were enriched in MCI. To analyze protein dynamics across disease progression, top DAPs for Aβ– and tau-PET were plotted against AD pseudotime which highlighted early increases for SMOC1 post-Aβ-positivity and early declines for Aβ42. NRGN and VSNL1 rose continuously, whereas MSLN, VEGFA and NPTX2 declined as pathology progressed. MDH1 and ENO2 increased gradually, with acceleration after tau-positivity (Fig. 4a).

**Figure 4:**
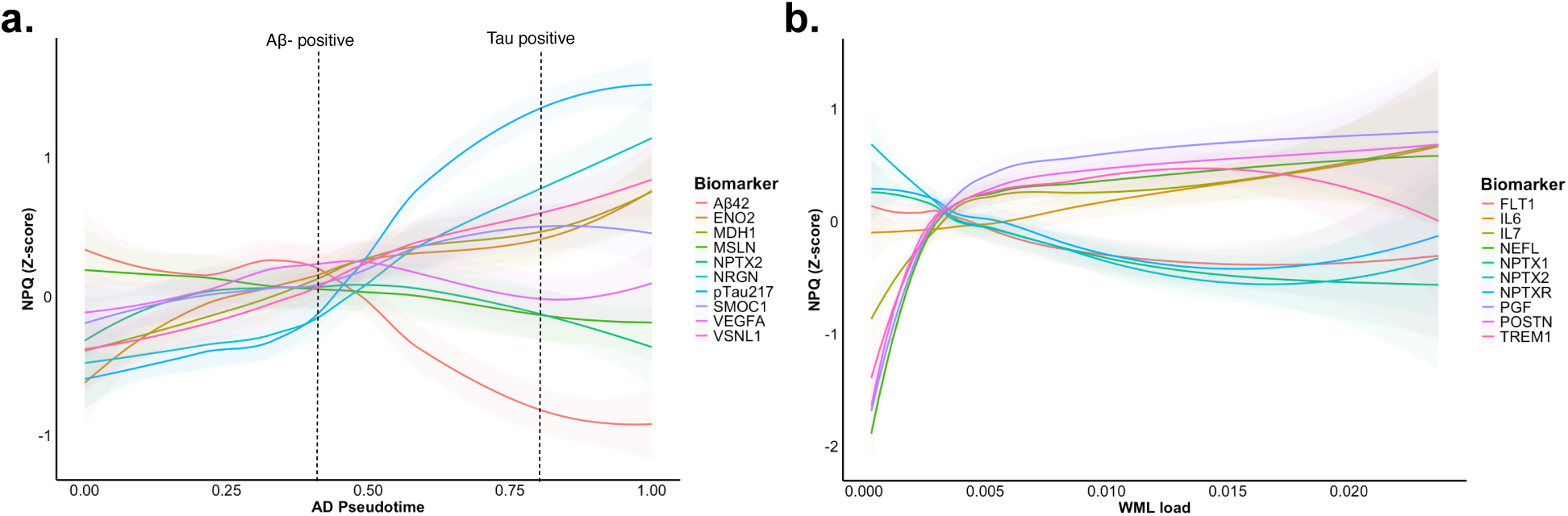
Differently abundant proteins levels across trajectories of disease. **a)** DAPs with the highest β standardized coefficients for AD fibrillary pathology are plotted against inferred AD pseudotime, with generalized additive model (GAM) lines. AD pseudotime error bands represent the 95% confidence interval (CI). Vertical dashed lines indicate positivity cutoffs: CSF Aβ42/Aβ40 Lumipulse G < 0.072 for Aβ and tau-PET Braak I-IV SUVR > 1.36 for tau. **b)** DAPs with highest β standardized coefficients for WML are plotted against WML load Vertical dashed line indicates the positivity cutoff for vascular pathology (WML % of ICV = 0.00594). Error bands represent the 95% confidence interval (CI).

The WML load profile was distinct, with 24 exclusive DAPs (Fig. 3b). FLT1, IL7, CCL3, CCL11, GOT1, SOD1, NEFL, and POSTN were the ones significant in both CU and MCI. Some of the main proteins in relation to WML are shown in Fig. 4b with similar trajectories across proteins with positive (PGF, POSTN, TREM1, IL16 and IL17; 0.23< std β < 0.13, p < 0.001) and negative associations (NPTX1, NPTX2, NPTXR, and FLT1; (−0.22 < std β< –0.14, p < 0.001).

### 5. Associations with longitudinal changes in pathology

Subsequently, we investigated whether any previously identified CSF DAPs were associated with the rate of change in Aβ-PET, tau-PET, or WML load. α-Synuclein was not included as only cross-sectional measures were available. We included 656 BioFINDER-2 participants with longitudinal data available for all three pathologies (Supplemental Table 4). We also performed separate analyses in CU and MCI groups to assess potential stage-specific effects. Protein associations with longitudinal changes in pathology are summarized in Supplemental Figure 6.

Baseline levels of four proteins (MAPT, MDH1, NRGN and VSNL1) were associated with worse progression of all three brain pathologies (Fig. 5a), positively with Aβ– and tau-PET and negatively with WML. VSNL1 was the only protein associated with the progression of the three pathologies in CU and MCI (Supplemental Fig. 7). Overall, a larger number of baseline proteins were associated with worsening of tau and WML pathology. Baseline NPTXR and NPTX2 levels were negatively, and NFL positively, associated with both tau-PET change and WML progression, whereas PGF, IL6, and ENO2 showed opposite associations with tau-PET change and WML progression (Fig. 5a). Sensitivity analyses showed that independent models for each pathology yielded more significant protein hits and overlap (Supplemental Fig. 8).

**Figure 5:**
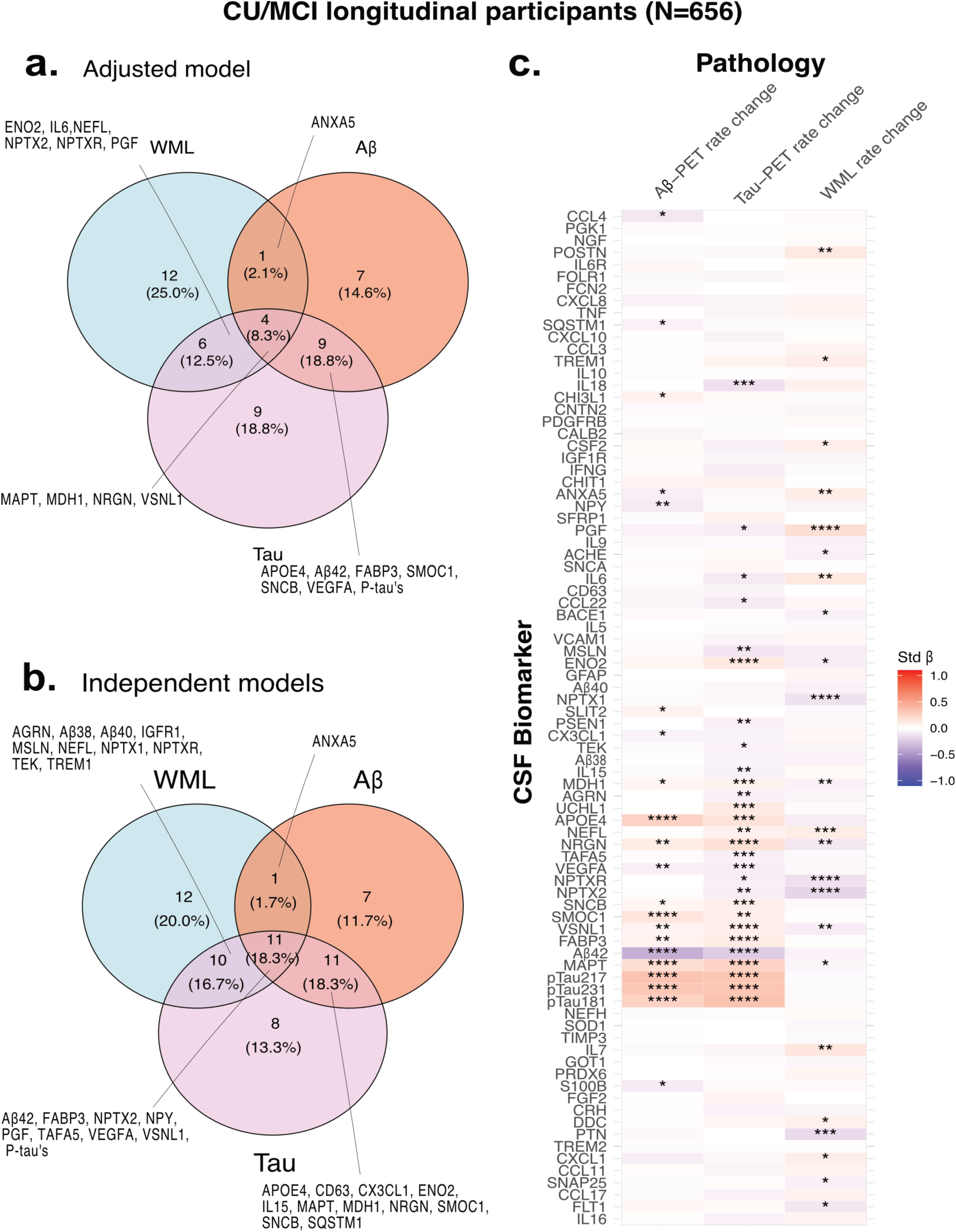
Associations between baseline DAPs and longitudinal changes in pathology. **Panels a-b** show Venn diagrams of DAPs associated with changes in amyloid, tau and WML pathology. **(a)** displays results from a single mutually adjusted model including all three pathologies, whereas **(b)** shows results from independent models for each pathology. **Panel c** shows a heatmap of standardized beta coefficients from the mutually adjusted model. Rows correspond to DAPs and columns to each pathology. Cell color reflects the direction (red for positive, blue for negative association) and strength (intensity of color proportional to absolute beta value) of each association between protein concentration and pathology burden. All models were adjusted for age, sex, and CSF protein levels. Analysis focused on previously identified DAPs. \**P_FDR_* < 0.05, *\*\*P_FDR_* < 0.01, *\*\*\*P_FDR_* < 0.001, *\*\*\*\*P_FDR_* < 0.0001.

The baseline levels of a core set of proteins were associated with worse progression of both Aβ and tau pathology, namely p-tau species, FABP3, SMOC1, SNCB, and APOE4 (0.05 < std β < 0.33, p < 0.04), whereas Aβ42 and VEGFA showed negative associations (−0.37 < std β< –0.08, p < 0.005) (Fig. 5c). Proteins uniquely associated with change in Aβ-PET included SLIT2, CHI3L1 (std β= 0.07, p<0.01), S100B, CCL4, NPY, SQSTM1and CX3CL1 and (−0.11< std β < –0.06, p < 0.01). Tau-PET rate of change was specifically associated with UCHL1 (std β= 0.12, p<0.001), MSLN, IL18, CCL22, IL15, TAFA5, PSEN1, AGRN, and TEK (−0.14 < std β < –0.07, p < 0.02). Stratifying participants by baseline cognitive stage revealed stage-specific differences (Supplemental Fig. 7). In CU, proteins associated with Aβ-specific progression included SMOC1, APOE4, SLIT2, CD63, and S100B while proteins associated with tau progression included ENO2, UCHL1, SNCB, and TAFA5. In MCI, no protein was uniquely linked to Aβ progression. In MCI, more associations were seen with tau-PET progression, including ENO2, MSLN, FABP3, APOE4, VSNL1, IL15, IL18, and PSEN1. These results indicate a broader, amyloid-linked proteomic response in the preclinical stage that narrows into a tau-centric profile in symptomatic stages.

WML rate of change was largely distinct, with 12 exclusive hits (Fig. 5c). Across cognitive stages, baseline levels of IL7 and PGF (std β= 0.15 and 0.18, p<0.001, respectively), and neuropentraxins (−0.19 < std β< –0.13, p < 0.001) were consistently associated with WML change. CU-specific associations included FLT1, PTN and MDH1, whereas POSTN, BACE1, and SNCB characterized WML progression in MCI.

### 6. Associations with brain atrophy and cognitive decline

Lastly, we examined whether any protein levels could predict brain atrophy or cognitive decline beyond baseline pathology measures (i.e., beyond Aβ– and tau-PET SUVR, WML load, and the presence of α-synuclein pathology). Longitudinal data were available for brain atrophy (n=914) and cognitive performance (n=1054). After accounting for baseline pathological burden, only a limited number of proteins contributed significantly to predicting cortical atrophy (Fig. 6a) or cognitive decline (Fig. 6b).

**Figure 6:**
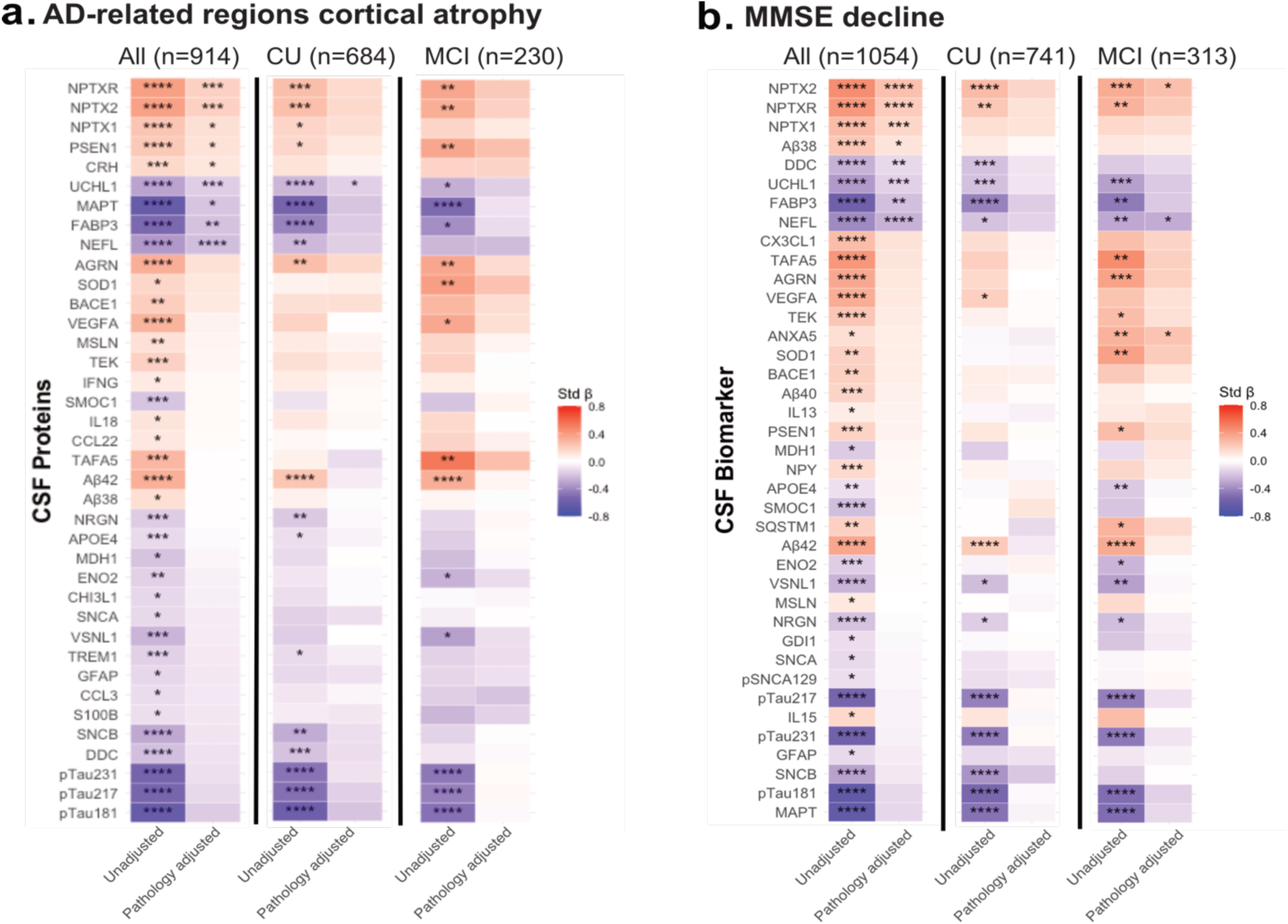
Prediction of AD-signature cortical atrophy and cognitive decline by baseline DAPs in BioFINDER-2. **Panel a** shows heatmaps of standardized beta coefficients for the association between baseline DAP levels and annualized atrophy rate in AD-signature cortical regions. **Panel b** presents analogous heatmaps for prediction of longitudinal change in Mini-Mental State Examination (MMSE) score. For each outcome, three heatmaps are displayed side-by-side: all participants, cognitively unimpaired (CU), and mild cognitive impairment (MCI). Within each heatmap, rows represent DAPs and the two columns correspond to the unadjusted protein association (left) and the association adjusted for baseline pathology burden (right). Cell color indicates direction (red for positive, blue for negative association) and strength (color intensity proportional to standardized beta value), with grey denoting non-significance. All models are adjusted for age, sex, and CSF protein levels, with additional adjustment for education in models of cognitive decline. \**P_FDR_* < 0.05, *\*\*P_FDR_* < 0.01, *\*\*\*P_FDR_* < 0.001, *\*\*\*\*P_FDR_* < 0.0001.

Higher concentration of UCHL1, NEFL MAPT, FABP3 were associated with greater cortical thinning within AD-signature regions (−0.22 < std β< –0.17, p < 0.001), whereas lower levels of PSEN1, CRH and neuropentraxins were associated with greater neurodegeneration in AD-signature regions (0.10< std β < 0.18, p < 0.001). Consistent associations were also observed when examining whole-brain volume reduction (Supplemental Fig. 9a). In CU individuals, UCHL1 was the only protein that significantly predicted atrophy; no proteins were significantly associated among participants with MCI. These were similar proteins with those identified as predictors of AD-signature regions cortical thickness cross-sectionally (Supplemental Fig.10a)

Similar patterns were observed for longitudinal changes in cognitive function (MMSE scores, Fig. 6b; or mPACC scores, Supplemental Fig. 9b). Higher CSF levels of UCHL1, NEFL, FABP3, DDC, and CCL2 were associated with greater decline in MMSE scores (−0.26 < std β< –0.13, p < 0.004), whereas lower concentrations of Aβ38 and neuropentraxins were linked to greater decline in cognitive function (0.11< std β < 0.25, p < 0.04). In subgroup analyses, no proteins were significantly associated with cognitive decline among CU participants, while in the MCI group, NPTX2, ANXA5, and NEFL remained significant predictors. NPTX2 was specifically associated in CU individuals when assessing mPACC decline (Supplemental Fig. 9b). These overlapped with those identified as predictors of MMSE scores in the cross-sectional analyses (Supplemental Fig.10b)

## Discussion

In this study, we leveraged the NULISA platform to comprehensively measure CSF biomarker candidates potentially related to AD, vascular, and α-synuclein pathologies from the BioFINDER study. By employing mutually adjusted models that account for multiple underlying pathologies, we addressed the frequent co-occurrence, as observed in neuropathological studies (4, 5), that can obscure the distinct biological features of each condition. This analytic strategy enabled the identification of both shared and pathology-specific biomarker signatures and the monitoring of how their expression patterns changed with increasing pathology burden. Notably, only a small subset of proteins, including neuropentraxins (NPTXs) and UCHL1, were consistently dysregulated across all pathologies, suggesting that they reflect neurodegenerative processes common to different etiologies. In contrast, distinct protein profiles delineated each pathology, many of which exhibited stage and temporal specificity, differing between CU and MCI individuals and/or between cross-sectional and longitudinal analyses.

In cross-sectional analyses, Aβ and tau pathology showed only partly overlapping biomarker signatures. Aβ-PET burden related mainly to glial activation and inflammatory markers (CHIT1, CX3CL1, CD63, TREM1, S100B, TAFA5, IL5), consistent with neuroinflammatory processes. Several of these proteins were also linked to vascular pathology, suggesting neurovascular-amyloid interactions. These observations align with previous CSF proteomic studies associating Aβ-PET with chemokine signaling (13), reports of early decreases in plasticity-related proteins (NPY, SQSTM1) (12), and the identification of SMOC1 as an early marker of AD progression and Aβ (11, 40). In contrast, tau pathology displayed fewer associated proteins, primarily involved in metabolic and synaptic function (VSLN1, ACHE, ENO2 and MDH1) reflecting neuronal vulnerability and stress and consistent with reports linking tau-PET to glucose/pyruvate metabolism and protein degradation pathway (13). Many of these findings align to NULISA prior studies on CSF (35).

Many proteins were negatively associated with tau pathology but positively associated with WML burden, indicating potentially divergent mechanisms, as for example neuroinflammatory markers (IL6, IL18, IL16, IL15, CCL17, CSF2). WML pathology showed a distinct angiogenic, inflammatory and extracellular matrix remodeling-related profile (VCAM1, FLT1, POSTN, PGF, NEFL, GFAP), with limited overlap beyond shared immune and degeneration pathways and consistent with blood-brain barrier breakdown (41). In contrast, α-synucleinopathy showed a more limited change, with significantly increased CSF DDC concentration, consistent with prior findings (42, 43).

Furthermore, the temporal dynamics of these biomarker signatures revealed clear stage-dependent patterns. In CU individuals, more DAPs were linked to Aβ burden, reflecting early glial and immune changes, as well as neurodegeneration, before clinical symptoms emerge. In contrast, in MCI individuals, tau biomarker changes predominated, consistent with tau-related synaptic and neuronal injury. Higher baseline protein concentrations were associated with more widespread longitudinal tau-PET changes than with Aβ-PET changes, particularly with markers of neuronal injury, inflammation, and metabolism, highlighting the central role of tau in disease progression particularly in symptomatic stages (44, 45). Interestingly, we identified MSLN, a cell-adhesion glycoprotein, as one of the most strongly associated with tau-PET increase, particularly in MCI, which had not been reported before to the best of our knowledge. The overlap between proteins associated with tau-PET change and WML progression was more pronounced than that between Aβ-PET change and WML progression, and predominantly involved synaptic and neuronal proteins (NRGN, VSNL1, NPTX2, NPTXR, NEFL, ENO2, MDH1) that were shared between both brain pathologies. This pattern is consistent with longitudinal AD proteomic trajectories showing persistent loss of synapses and neurons across the disease continuum (12) and aligns with evidence that white matter injury can also contribute to neuronal signaling deficits (15, 46). This likely suggests that AD and WML pathologies, although initiated by distinct mechanisms, may converge on shared downstream pathways that disrupt neuronal signaling and culminate in synaptic and neuronal loss. Interestingly, proteins that predicted cognitive decline and brain atrophy showed remarkable consistency, with most being previously identified as shared across multiple neurodegenerative pathologies (neuropentraxins, FABP3, UCHL1 and Aβ38) (12, 47), as well as NEFL, a known marker of general degeneration (7). Their central involvement across disease mechanisms likely enables them to capture core processes driving cognitive decline and brain atrophy.

Plasma results were less sensitive, detecting mainly established markers of AD (p-tau217, p-tau181, p-tau231, GFAP, Aβ42) in line with previous results (48–50). While others using NULISA identified plasma proteins such as NRGN, CHIT3, BDNF, ANXA5, CCL2, SOD1, and NPY to be related to Aβ-status or clinical AD diagnosis, our stricter definition requiring both Aβ– and tau-PET positivity and also adjusting for other pathologies explains their absence (49, 51, 52). Larger cohorts using NULISA similarly reported PSEN1, ACHE and BDNF to be changed selectively in AD (50), while NEFL consistently elevated across different neurodegenerative diseases (35, 48–50). Overall, while plasma NULISA appears useful for detecting AD-related changes, CSF emerges as the more informative matrix for capturing multi-pathology effects and assessing shared molecular signatures.

### Limitations

Our classification approach focused on individuals with established pathology, which may have limited detection of earlier proteomic changes. The binary classification of α-synucleinopathy by RT-QuIC captures the presence of pathology but not its severity. Interaction effects between pathologies were not explicitly modeled potentially missing additive or synergistic effects. Cerebral small vessel disease does not only encompass WMLs, but also lacunar infarcts and microbleeds. Still, this study focused solely on WML, since it is the feature that is most common and consistently associated with clinical outcomes. Plasma analyses, while informative, remain less sensitive for detecting co-occurring non-AD processes, which highlights the need for unsupervised discovery and multiplexed platforms to capture non-AD signatures. The predominance of white individuals in our cohort may restrict the generalizability of these findings. Finally, as classifications were based on in vivo biomarkers, neuropathological validation will be important; future studies integrating pre-mortem CSF/plasma with postmortem brain data are needed to refine disease-specific proteomic signatures.

## Conclusion

In conclusion, the NULISA platform identified distinct protein signatures for AD, α-synucleinopathy, and vascular pathology, with limited overlap across conditions. These signatures showed stage-dependent shifts aligned with pathology burden, highlighting early glial and inflammatory responses to Aβ, progressive synaptic and neuronal injury linked to tau, and largely independent angiogenic and structural remodeling related to white matter integrity. Shared proteins, including neuropentraxins, UCHL1, and NEFL, reflect convergent neurodegenerative processes underlying cognitive decline and atrophy. Together, these results emphasize that integrating shared and pathology-specific dynamics through CSF proteomics can refine biomarker stratification and improve understanding of neurodegenerative disease progression.

## Declarations

### Ethics approval

The ethical approval was given by the Regional Ethics Committee in Lund, Sweden. All participants gave their written informed consent to participate in the study, and study procedures were conducted in accordance with the Declaration of Helsinki.

### Consent for publication

Not applicable

### Availability of data and material

Pseudonymized data will be shared upon request from a qualified academic investigator for the purpose of replicating procedures and results presented in the study. For this procedure a data transfer agreement must be established with Skåne University Hospital (Region Skåne) to share data in agreement with EU legislation. The proposed analyses must be compliant with decisions made by the Swedish Ethical Review Authority.

### Competing interests

GS has received speaker fees from Springer, GE Healthcare, Biogen, Esteve and Adium and she serves on the scientific advisory board of Johnson&Johnson. SP has acquired research support (for the institution) from Avid Radiopharmaceuticals and ki elements through ADDF. In the past 3 years, he has received consultancy/speaker fees from BioArtic, Danaher, Eisai, Eli Lilly, Novo Nordisk, and Roche. HZ has served at scientific advisory boards and/or as a consultant for Abbvie, Acumen, Alector, Alzinova, ALZPath, Amylyx, Annexon, Apellis, Artery Therapeutics, AZTherapies, Cognito Therapeutics, CogRx, Denali, Eisai, LabCorp, Merry Life, Nervgen, Novo Nordisk, Optoceutics, Passage Bio, Pinteon Therapeutics, Prothena, Red Abbey Labs, reMYND, Roche, Samumed, Siemens Healthineers, Triplet Therapeutics, and Wave, has given lectures in symposia sponsored by Alzecure, Biogen, Cellectricon, Fujirebio, Lilly, Novo Nordisk, and Roche, and is a co-founder of Brain Biomarker Solutions in Gothenburg AB (BBS), which is a part of the GU Ventures Incubator Program (outside submitted work). NJA has served as consultant for Quanterix and has given lectures in symposia sponsored by Lilly, Quanterix and BiogenNMC has received consultancy/speaker fees from Biogen, BioArctic, Lilly, Merck, Novo Nordisk, and Owkin. OH, is an employee of Lund University and Lilly. All other authors report no competing interest.

## Funding information

Work in the authors’ research group was supported by the National Institute of Aging (R01AG083740), the Alzheimer’s Association, (22AARF-1029663, ZEN24-1069572) the Alzheimer’s Association in collaboration with the GHR Foundation (ALZSI-26-1523522), the European Research Council (ADG-101096455 and ADG-101053962), European Commission/Horizon Europe (101108819, 101153323), ERA PerMed (ERAPERMED2021-184), The Michael J Fox Foundation (MJFF-025507, MJFF-025741), Global Research Platform LLC, the Cure Alzheimer’s fund (F2025/1066), Swedish Research Council (2021-02219, 2022-00775, 2023-06428), the Swedish Alzheimer Foundation (AF-981132, AF-980907 and AF-968598, AF-1011949, AF-1011799), the Swedish Brain Foundation (FO2023-0163, FO2024-0133-HK-46, FO2024-0284,FO2024-0385, FO2025-0055), The Swedish Parkinson Foundation (1485/23, 1589/24), the Strategic Research Area MultiPark (Multidisciplinary Research in Parkinson’s disease) at Lund University, WASP and DDLS Joint call for research projects (WASP/DDLS22-066), the Skåne University Hospital Foundation, Regional research support (2025-2024-2925, 2025-2024-2862), the Swedish federal government under the ALF agreement (2022-Project0080, 2022-Project0107, 2022-Project0062, 2022-YF0017, 2024-YF0048, 2024-ST0026, ALFGBG-71320), Lilly Research Award Program (F2025/932), Avid Pharmaceuticals, F Hoffman-la Roche AG (F2025/453), Biogen, Bristol Myers Squibb, CellInvent (F2025/1321), Eisai, Fujirebio, GE Healthcare, the Knut and Alice Wallenberg foundation (2017-0383), Konung Gustaf V:s och Drottning Victorias Frimurarestiftelse, the Kamprad Foundation (20243058), Bundy Academy, Greta and Johan Kock Foundation (F2024/228, F2024/2330, F2024/197, F2025/160, F2024/198, F2024/2332, F2025/319), the Crafoord Foundation (20240888), the Rönström Family Foundation (AF1011799, FRS 0013, FRS-0003, FRS-0004, AF-1011949, the Royal Physiographic Society (F2025/1092), Stiftelsen för Gamla Tjänarinnor (2024-250), Thorsten and Elsa Segerfalk’s FoundationF2024/1053, F2025/1320), and Fredrik and Ingrid Thuring’s Foundation (2024-099), the Alzheimer Drug Discovery Foundation (ADDF), USA (#201809-2016862), the AD Strategic Fund and the Alzheimer’s Association (#ADSF-21-831376-C, #ADSF-21-831381-C, #ADSF-21-831377-C, and #ADSF-24-1284328-C), and the European Partnership on Metrology, co-financed from the European Union’s Horizon Europe Research and Innovation Programme and by the Participating States (NEuroBioStand, #22HLT07).The funding sources had no role in the design and conduct of the study; in the collection, analysis, interpretation of the data; or in the preparation, review, or approval of the manuscript.

## Authors contribution

AOD, APB, GS, SJ, NMC and OH contributed to the study’s conception and design. SP and ES contributed with data collection.ALB, IP, KT, WT, HZ, NJA contributed with NULISA data. AM and PP contributed with SAA data. AOD performed data analysis and produced the figures. AOD and APB were major contributors in writing the manuscript. All authors reviewed the manuscript and provided feedback. SP, NMC, OH secured funding for the present study.

## Abbreviations

ADAS-Cog: Alzheimer’s Disease Assessment Scale-Cognitive Subscale
AD: Alzheimer’s disease
Aβ: amyloid-β
BP: biological process
cSVD: cerebral small vessel disease
CNS: Central nervous system
CSF: cerebrospinal fluid
CU: cognitively unimpaired
DAP: Differentially abundant proteins
EDTA: Ethylenediaminetetraacetic acid
FDR: false discovery rate
FLAIR: fluid-attenuated inversion recovery
GAM: generalized additive model
GO: gene ontology
ICV: intracranial volume
IQR: interquartile range
LBD: Lewy Body Disease
LOD: limit of detection
MCI: mild cognitive impaired
MMSE: Mini-Mental State Examination
MRI: magnetic resonance imaging
MPRAGE: magnetization-prepared rapid gradient echo
MTG: middle temporal gyrus
NPQ: NULISA Protein Quantification
NULISA: Nucleic Acid Linked Immuno-Sandwich Assay
OPCs: oligodendrocyte precursor cells
ACC: Preclinical Alzheimer’s Cognitive Composite
PD: Parkinson’s disease
PET: positron emission transmission
RT-QuIC: Real-Time Quaking-Induced Conversion
SAA: seed amplification assay
SAMSEG: Sequence Adaptive Multimodal SEGmentation
SCD: subjective cognitive decline
SD: standard deviation
SUVR: Standardized uptake value ratios
VaD: Vascular Disease
WebGestalt: WEB-based Gene SeTAnaLysis Toolkit
WMH: white matter hyperintensities
WML: white matter lesion

## Data Availability

Pseudonymized data will be shared upon request from a qualified academic investigator for the purpose of replicating procedures and results presented in the study. For this procedure a data transfer agreement must be established with Skane University Hospital (Region Skane) to share data in agreement with EU legislation. The proposed analyses must be compliant with decisions made by the Swedish Ethical Review Authority.

## Supplementary material

Supplementary material is available at *Molecular Neurodegeneration* online.

## FIG

## Notes

### Clinical Trial

NCT01208675; NCT03174938

## References

1. Rahimi J, Kovacs GG. Prevalence of mixed pathologies in the aging brain. Alzheimers Res Ther. 2014;6(9):82.

2. Kovacs GG, Milenkovic I, Wohrer A, Hoftberger R, Gelpi E, Haberler C, et al. Non-Alzheimer neurodegenerative pathologies and their combinations are more frequent than commonly believed in the elderly brain: a community-based autopsy series. Acta Neuropathol. 2013;126(3):365–84.

3. Kapasi A, DeCarli C, Schneider JA. Impact of multiple pathologies on the threshold for clinically overt dementia. Acta Neuropathol. 2017;134(2):171–86.

4. Schneider JA, Arvanitakis Z, Bang W, Bennett DA. Mixed brain pathologies account for most dementia cases in community-dwelling older persons. Neurology. 2007;69(24):2197–204.

5. Deramecourt V, Slade JY, Oakley AE, Perry RH, Ince PG, Maurage CA, Kalaria RN. Staging and natural history of cerebrovascular pathology in dementia. Neurology. 2012;78(14):1043–50.

6. Palmqvist S, Rossi M, Hall S, Quadalti C, Mattsson-Carlgren N, Dellavalle S, et al. Cognitive effects of Lewy body pathology in clinically unimpaired individuals. Nat Med. 2023;29(8):1971–8.

7. Hansson O. Biomarkers for neurodegenerative diseases. Nat Med. 2021;27(6):954–63.

8. Jack CR, Jr., Andrews JS, Beach TG, Buracchio T, Dunn B, Graf A, et al. Revised criteria for diagnosis and staging of Alzheimer’s disease: Alzheimer’s Association Workgroup. Alzheimers Dement. 2024;20(8):5143–69.

9. Jack CR, Jr., Bennett DA, Blennow K, Carrillo MC, Dunn B, Haeberlein SB, et al. NIA-AA Research Framework: Toward a biological definition of Alzheimer’s disease. Alzheimers Dement. 2018;14(4):535–62.

10. Hansson O, Edelmayer RM, Boxer AL, Carrillo MC, Mielke MM, Rabinovici GD, et al. The Alzheimer’s Association appropriate use recommendations for blood biomarkers in Alzheimer’s disease. Alzheimers Dement. 2022;18(12):2669–86.

11. Pichet Binette A, Gaiteri C, Wennstrom M, Kumar A, Hristovska I, Spotorno N, et al. Proteomic changes in Alzheimer disease associated with progressive Abeta plaque and tau tangle pathologies. Nat Neurosci. 2024.

12. Ali M, Timsina J, Western D, Liu M, Beric A, Budde J, et al. Multi-cohort cerebrospinal fluid proteomics identifies robust molecular signatures across the Alzheimer disease continuum. Neuron. 2025.

13. Wang Z, Chen Y, Gong K, Zhao B, Ning Y, Chen M, et al. Cerebrospinal fluid proteomics identification of biomarkers for amyloid and tau PET stages. Cell Rep Med. 2025:102031.

14. Weiner S, Sauer M, Montoliu-Gaya L, Benedet AL, Ashton NJ, Gonzalez-Ortiz F, et al. Cerebrospinal fluid proteome profiling across the Alzheimer’s disease continuum: a step towards solving the equation for ‘X’. Mol Neurodegener. 2025;20(1):52.

15. Hristovska I, Binette AP, Kumar A, Gaiteri C, Karlsson L, Strandberg O, et al. Identification of distinct and shared biomarker panels in different manifestations of cerebral small vessel disease through proteomic profiling. medRxiv. 2024.

16. Dammer EB, Ping L, Duong DM, Modeste ES, Seyfried NT, Lah JJ, et al. Multi-platform proteomic analysis of Alzheimer’s disease cerebrospinal fluid and plasma reveals network biomarkers associated with proteostasis and the matrisome. Alzheimers Res Ther. 2022;14(1):174.

17. Feng W, Beer JC, Hao Q, Ariyapala IS, Sahajan A, Komarov A, et al. NULISA: a proteomic liquid biopsy platform with attomolar sensitivity and high multiplexing. Nat Commun. 2023;14(1):7238.

18. Petrazzuoli F, Vestberg S, Midlov P, Thulesius H, Stomrud E, Palmqvist S. Brief Cognitive Tests Used in Primary Care Cannot Accurately Differentiate Mild Cognitive Impairment from Subjective Cognitive Decline. J Alzheimers Dis. 2020;75(4):1191–201.

19. American Psychiatric Association., American Psychiatric Association. DSM-5 Task Force. Diagnostic and statistical manual of mental disorders: DSM-5. 5th ed. Washington, D.C.: American Psychiatric Association; 2013. xliv, 947 p. p.

20. Dubois B, Villain N, Schneider L, Fox N, Campbell N, Galasko D, et al. Alzheimer Disease as a Clinical-Biological Construct-An International Working Group Recommendation. JAMA Neurol. 2024;81(12):1304–11.

21. Palmqvist S, Janelidze S, Quiroz YT, Zetterberg H, Lopera F, Stomrud E, et al. Discriminative Accuracy of Plasma Phospho-tau217 for Alzheimer Disease vs Other Neurodegenerative Disorders. JAMA. 2020;324(8):772–81.

22. Papp KV, Rentz DM, Orlovsky I, Sperling RA, Mormino EC. Optimizing the preclinical Alzheimer’s cognitive composite with semantic processing: The PACC5. Alzheimers Dement (N Y). 2017;3(4):668–77.

23. Ossenkoppele R, Pichet Binette A, Groot C, Smith R, Strandberg O, Palmqvist S, et al. Amyloid and tau PET-positive cognitively unimpaired individuals are at high risk for future cognitive decline. Nat Med. 2022;28(11):2381–7.

24. Palmqvist S, Scholl M, Strandberg O, Mattsson N, Stomrud E, Zetterberg H, et al. Earliest accumulation of beta-amyloid occurs within the default-mode network and concurrently affects brain connectivity. Nat Commun. 2017;8(1):1214.

25. Cerri S, Puonti O, Meier DS, Wuerfel J, Muhlau M, Siebner HR, Van Leemput K. A contrast-adaptive method for simultaneous whole-brain and lesion segmentation in multiple sclerosis. Neuroimage. 2021;225:117471.

26. Bronte A, Prieto E, Quincoces G, Erro E, Arbizu J. The basics of PET molecular imaging in neurodegenerative disorders with dementia and/or parkinsonism. Eur Radiol. 2025.

27. Jack CR, Jr., Wiste HJ, Weigand SD, Therneau TM, Lowe VJ, Knopman DS, et al. Defining imaging biomarker cut points for brain aging and Alzheimer’s disease. Alzheimers Dement. 2017;13(3):205–16.

28. Leuzy A, Smith R, Ossenkoppele R, Santillo A, Borroni E, Klein G, et al. Diagnostic Performance of RO948 F 18 Tau Positron Emission Tomography in the Differentiation of Alzheimer Disease From Other Neurodegenerative Disorders. JAMA Neurol. 2020;77(8):955–65.

29. Blennow K, Hampel H, Weiner M, Zetterberg H. Cerebrospinal fluid and plasma biomarkers in Alzheimer disease. Nat Rev Neurol. 2010;6(3):131–44.

30. Gobom J, Parnetti L, Rosa-Neto P, Vyhnalek M, Gauthier S, Cataldi S, et al. Validation of the LUMIPULSE automated immunoassay for the measurement of core AD biomarkers in cerebrospinal fluid. Clin Chem Lab Med. 2022;60(2):207–19.

31. Orru CD, Ma TC, Hughson AG, Groveman BR, Srivastava A, Galasko D, et al. A rapid alpha-synuclein seed assay of Parkinson’s disease CSF panel shows high diagnostic accuracy. Ann Clin Transl Neurol. 2021;8(2):374–84.

32. Chao LL, Decarli C, Kriger S, Truran D, Zhang Y, Laxamana J, et al. Associations between white matter hyperintensities and beta amyloid on integrity of projection, association, and limbic fiber tracts measured with diffusion tensor MRI. PLoS One. 2013;8(6):e65175.

33. DeCarli C, Murphy DG, Tranh M, Grady CL, Haxby JV, Gillette JA, et al. The effect of white matter hyperintensity volume on brain structure, cognitive performance, and cerebral metabolism of glucose in 51 healthy adults. Neurology. 1995;45(11):2077–84.

34. Rossi M, Candelise N, Baiardi S, Capellari S, Giannini G, Orru CD, et al. Ultrasensitive RT-QuIC assay with high sensitivity and specificity for Lewy body-associated synucleinopathies. Acta Neuropathol. 2020;140(1):49–62.

35. Ashton NJ, Benedet AL, Molfetta GD, Pola I, Anastasi F, Fernandez-Lebrero A, et al. Biomarker discovery in Alzheimer’s and neurodegenerative diseases using Nucleic Acid Linked Immuno-Sandwich Assay. Alzheimers Dement. 2025;21(5):e14621.

36. Karlsson L, Vogel J, Arvidsson I, Astrom K, Janelidze S, Blennow K, et al. Cerebrospinal fluid reference proteins increase accuracy and interpretability of biomarkers for brain diseases. Nat Commun. 2024;15(1):3676.

37. Satija R, Farrell JA, Gennert D, Schier AF, Regev A. Spatial reconstruction of single-cell gene expression data. Nat Biotechnol. 2015;33(5):495–502.

38. Hao Y, Hao S, Andersen-Nissen E, Mauck WM, 3rd, Zheng S, Butler A, et al. Integrated analysis of multimodal single-cell data. Cell. 2021;184(13):3573–87 e29.

39. Elizarraras JM, Liao Y, Shi Z, Zhu Q, Pico AR, Zhang B. WebGestalt 2024: faster gene set analysis and new support for metabolomics and multi-omics. Nucleic Acids Res. 2024;52(W1):W415–W21.

40. Wang YT, Ashton NJ, Therriault J, Benedet AL, Macedo AC, Pola I, et al. Identify biological Alzheimer’s disease using a novel nucleic acid-linked protein immunoassay. Brain Commun. 2025;7(1):fcaf004.

41. Is O, Wang X, Reddy JS, Min Y, Yilmaz E, Bhattarai P, et al. Gliovascular transcriptional perturbations in Alzheimer’s disease reveal molecular mechanisms of blood brain barrier dysfunction. Nat Commun. 2024;15(1):4758.

42. Bolsewig K, Willemse EAJ, Sanchez-Juan P, Rabano A, Martinez M, Doecke JD, et al. Increased plasma DOPA decarboxylase levels in Lewy body disorders are driven by dopaminergic treatment. Nat Commun. 2025;16(1):1139.

43. Pereira JB, Kumar A, Hall S, Palmqvist S, Stomrud E, Bali D, et al. DOPA decarboxylase is an emerging biomarker for Parkinsonian disorders including preclinical Lewy body disease. Nat Aging. 2023;3(10):1201–9.

44. Salvado G, Ossenkoppele R, Ashton NJ, Beach TG, Serrano GE, Reiman EM, et al. Specific associations between plasma biomarkers and postmortem amyloid plaque and tau tangle loads. EMBO Mol Med. 2023;15(5):e17123.

45. La Joie R, Visani AV, Baker SL, Brown JA, Bourakova V, Cha J, et al. Prospective longitudinal atrophy in Alzheimer’s disease correlates with the intensity and topography of baseline tau-PET. Sci Transl Med. 2020;12(524).

46. Qin C, Yang S, Chu YH, Zhang H, Pang XW, Chen L, et al. Signaling pathways involved in ischemic stroke: molecular mechanisms and therapeutic interventions. Signal Transduct Target Ther. 2022;7(1):215.

47. Oh HS, Urey DY, Karlsson L, Zhu Z, Shen Y, Farinas A, et al. A cerebrospinal fluid synaptic protein biomarker for prediction of cognitive resilience versus decline in Alzheimer’s disease. Nat Med. 2025.

48. Rea Reyes RE, Wilson RE, Langhough RE, Studer RL, Jonaitis EM, Oomens JE, et al. Targeted proteomic biomarker profiling using NULISA in a cohort enriched with risk for Alzheimer’s disease and related dementias. Alzheimers Dement. 2025;21(5):e70166.

49. Durcan R, Heslegrave A, Swann P, Goddard J, Chouliaras L, Murley AG, et al. Novel blood-based proteomic signatures across multiple neurodegenerative diseases. Alzheimers Dement. 2025;21(3):e70116.

50. Gong K, Timsina J, Ali M, Chen Y, Liu M, Wang C, et al. High-sensitivity plasma proteomics reveals disease-specific signatures and predictive biomarkers of Alzheimer’s disease phenotypes in a large mixed-dementia cohort. Mol Neurodegener. 2025;20(1):120.

51. Zeng X, Lafferty TK, Sehrawat A, Chen Y, Ferreira PCL, Bellaver B, et al. Multi-analyte proteomic analysis identifies blood-based neuroinflammation, cerebrovascular and synaptic biomarkers in preclinical Alzheimer’s disease. Mol Neurodegener. 2024;19(1):68.

52. Zeng X, Sehrawat A, Lafferty TK, Chen Y, Rawat M, Kamboh MI, et al. Novel plasma biomarkers of amyloid plaque pathology and cortical thickness: Evaluation of the NULISA targeted proteomic platform in an ethnically diverse cohort. Alzheimers Dement. 2025;21(2):e14535.

