## Supplemental 1 for "Plasma and CSF proteomic signatures related to Alzheimer’s, α-synuclein, or vascular pathologies and clinical decline"

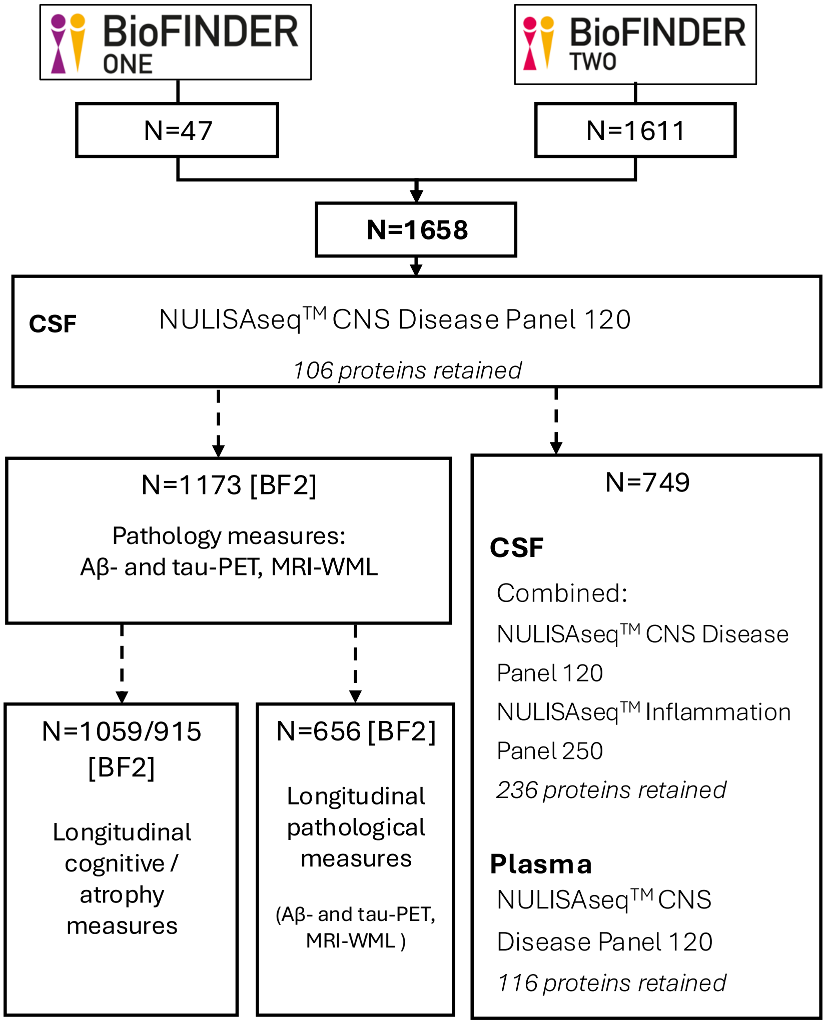


**Supplemental Figure 1: Overview of study design, participants, and proteomic analyses.** The figure summarizes the number of participants included in each analysis and sub-analysis, the biological samples assessed (CSF and/or plasma), and the corresponding NULISA proteomic panels (CNS Disease Panel and Inflammation Panel) applied.

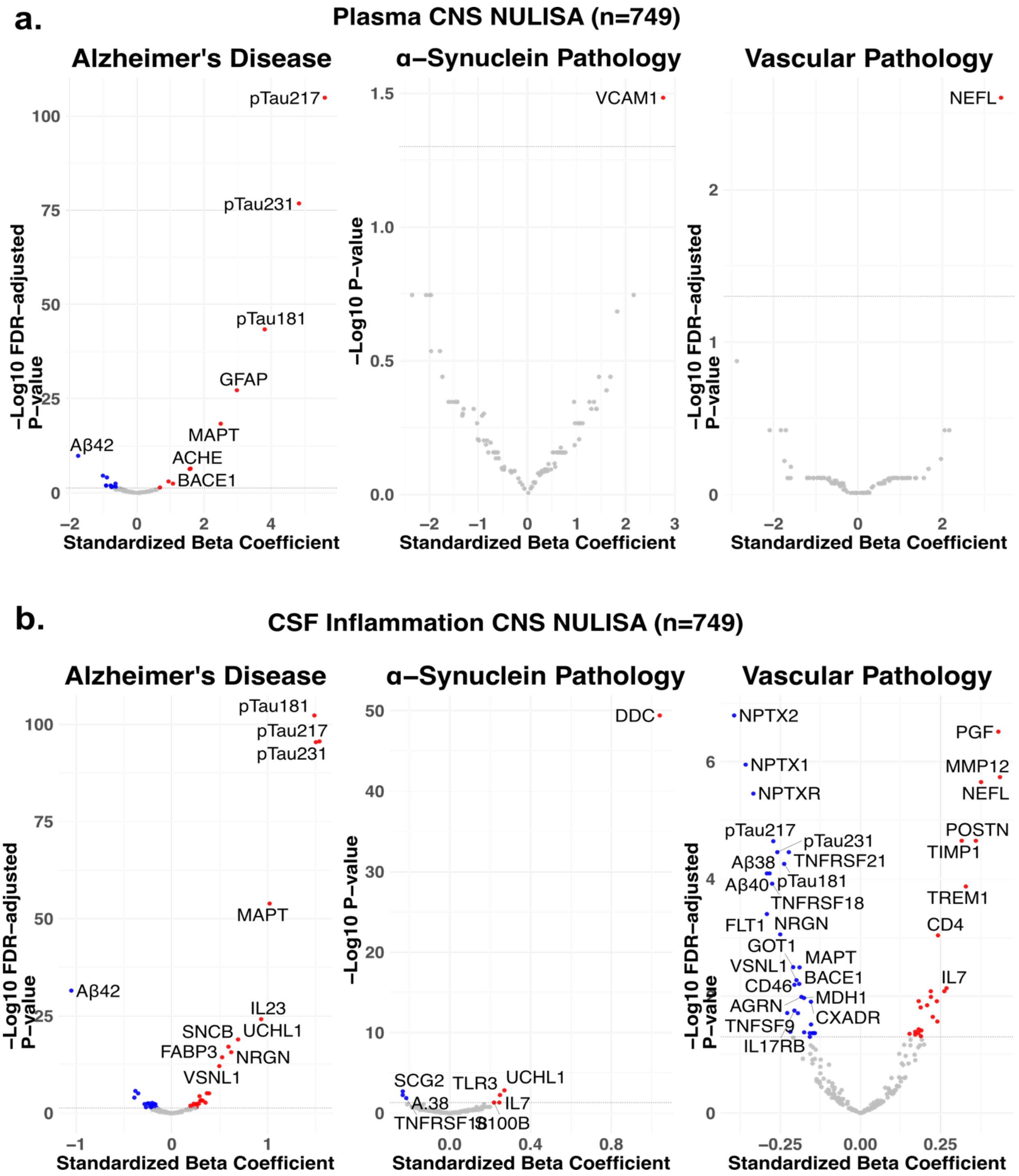


**Supplemental Figure 2: Identification of Differential Abundant Proteins across different diseases in a subcohort.**

Analysis in subcohort with both plasma CNS and CSF CNS and Inflammation data including 749 individuals**. a)** Volcano plots show proteins linked to AD, α-synuclein, and vascular pathology in plasma using CNS NULISA panel. The x-axis represents standardized β values (effect size), and the y-axis shows -log10 p-values (statistical significance). Model included all three pathology categories, age, sex, and CSF protein levels. Dashed lines mark significance at α=0.05 (FDR-corrected). Significant proteins (p[FDR]<0.05) are in red (higher abundance in presence of pathology) or blue (lower abundance). **b)** Volcano plots in CSF show proteins linked to AD, α-synuclein, and vascular pathology in CSF using CNS and Inflammation NULISA panel (n=324 proteins)

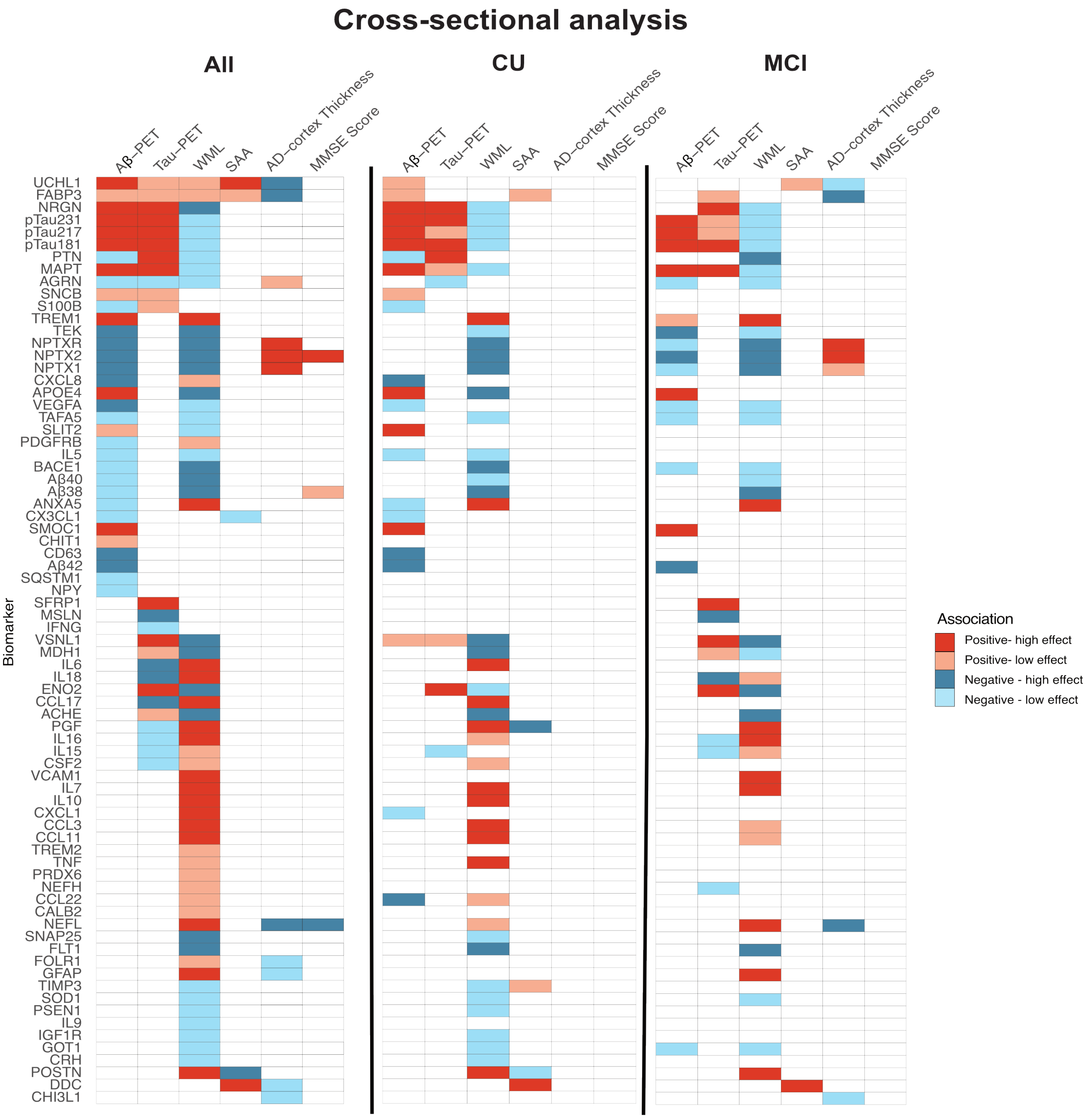

**Supplemental Figure 3: Baseline associations of DAPs with pathology biomarkers, cortical thickness measures, and cognitive scores across participant subgroups.**

Each tile represents the association between a biomarker and a specific feature (pathology, cortical thickness, or cognitive score), where tile color is feature-specific. Red tiles denote positive associations and blue tiles negative associations. More saturated shades mark proteins with larger effect sizes, while lighter shades mark smaller effect sizes. White tiles indicate no significant association. Results are displayed in separate panels for all participants (left), cognitively unimpaired (CU, center), and mild cognitive impairment (MCI, right) groups.

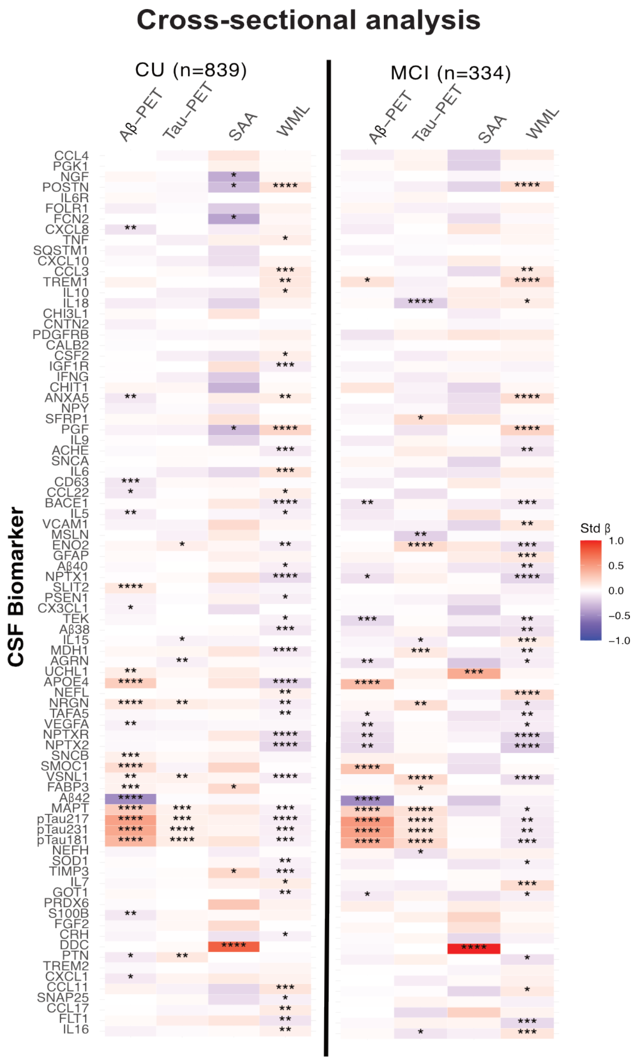


**Supplemental Figure 4: Associations between baseline concentrations of DAPs and load of pathology and presence of α-synuclein pathology**

Heatmaps illustrating standardized beta coefficients from the mutually adjusted model for all four pathologies, with associations shown separately for cognitively unimpaired (CU) participants (left) and participants with mild cognitive impairment (MCI) (right). Cell color reflects the direction (red for positive, blue for negative association) and strength (intensity of color proportional to absolute beta value) of each association between protein concentration and pathology burden. All models were adjusted for age, sex, and CSF protein levels. Analysis focused on previously identified DAPs. *$P_{FDR} < 0.05$, **$P_{FDR} < 0.01$, ***$P_{FDR} < 0.001$, ****$P_{FDR} < 0.0001$.

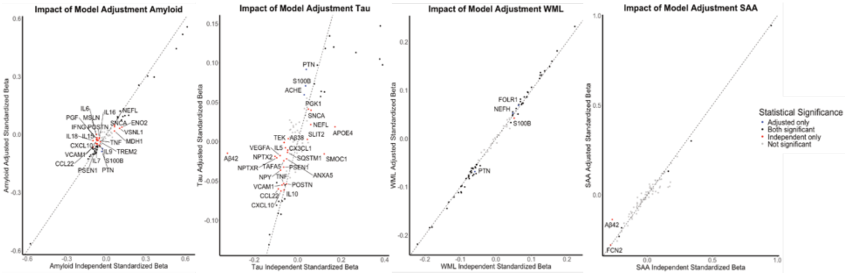


**Supplemental Figure 5: Comparison of standardized beta coefficients of DAPs identified using independent versus adjusted models associations to pathology load.**

Scatterplot showing proteins identified as significantly associated (FDR < 0.05) with a given pathology in either an independent model (x-axis) or an adjusted model accounting for all other pathologies (y-axis). Each point represents a protein, color-coded by significance pattern: black for proteins significant in both models, red for those significant in the independent model only, and blue for those significant in the adjusted model only. The dashed diagonal line indicates equal effect sizes across models.

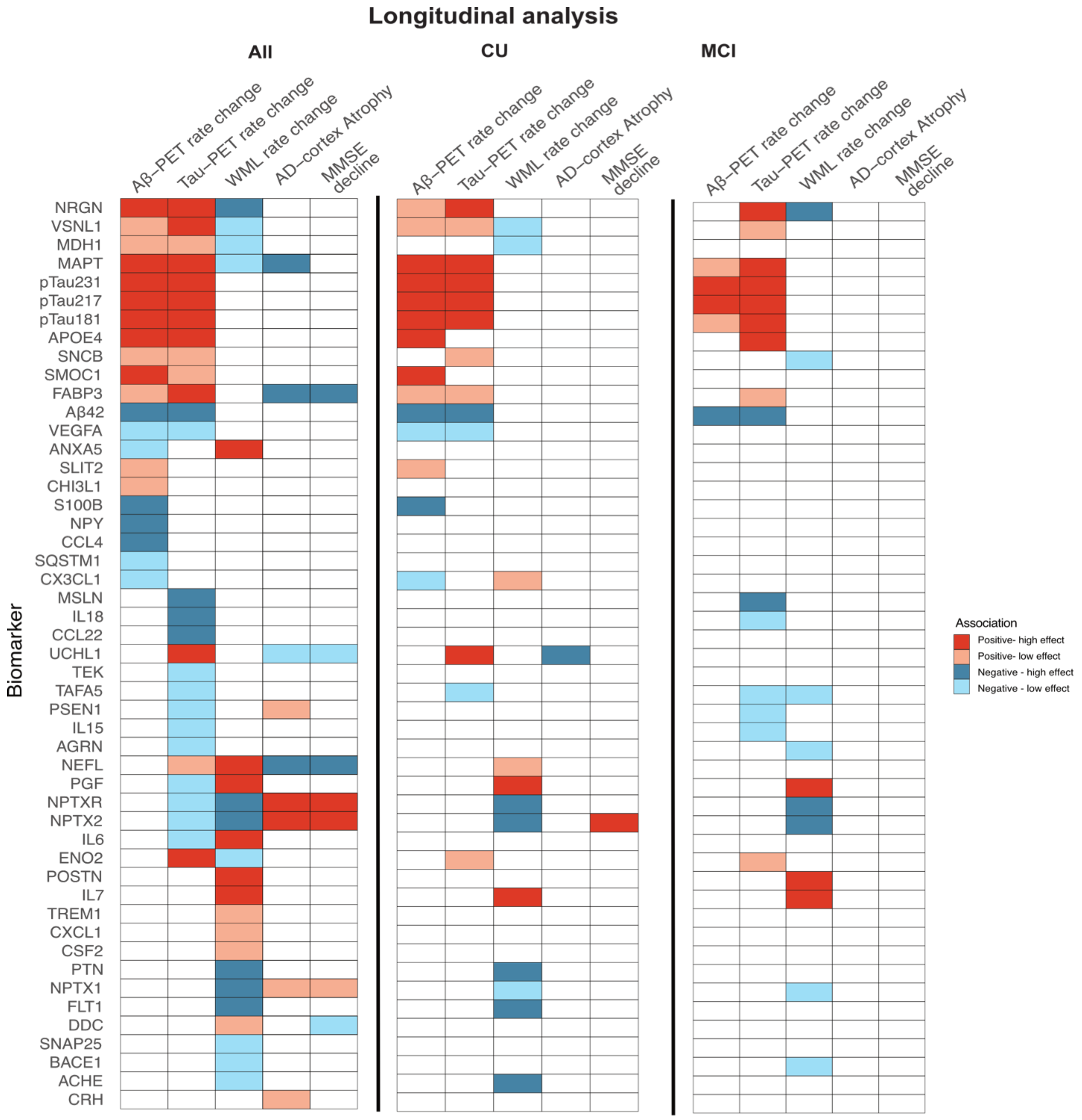


**Supplemental Figure 6: Associations between baseline levels of DAPs and subsequent longitudinal changes in pathology biomarkers (rate of change), brain atrophy measures, and cognitive decline across subgroups.**

Each tile depicts the association between a biomarker and a longitudinal feature. ed tiles denote positive associations and blue tiles negative associations. More saturated shades mark proteins with larger effect sizes, while lighter shades mark smaller effect sizes. White tiles indicate no significant association. Results are displayed in separate panels for all participants (left), cognitively unimpaired (CU, center), and mild cognitive impairment (MCI, right) groups.

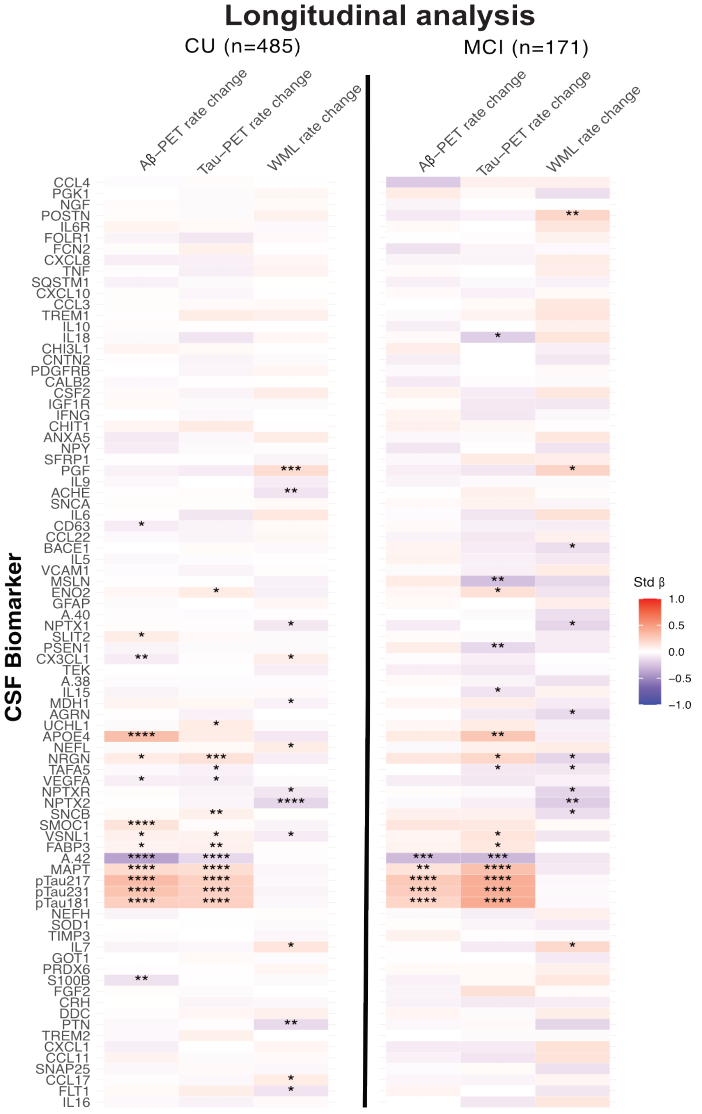


**Supplemental Figure 7: Associations between baseline concentrations of DAPs and longitudinal changes in pathology**

Heatmaps illustrating standardized beta coefficients from the mutually adjusted model, with associations shown separately for cognitively unimpaired (CU) participants (left) and participants with mild cognitive impairment (MCI) (right). Cell color reflects the direction (red for positive, blue for negative association) and strength (intensity of color proportional to absolute beta value) of each association between protein concentration and pathology burden. All models were adjusted for age, sex, and CSF protein levels. Analysis focused on previously identified DAPs. *$P_{FDR} < 0.05$, **$P_{FDR} < 0.01$, ***$P_{FDR} < 0.001$, ****$P_{FDR} < 0.0001$.

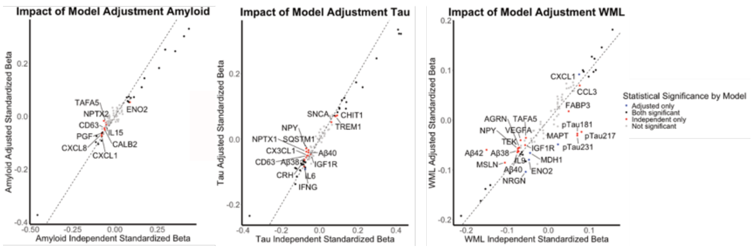


**Supplemental Figure 8: Comparison of standardized beta coefficients of DAPs using independent versus adjusted models associations with pathology rate of change**

Scatterplot showing proteins identified as significantly associated (FDR < 0.05) with a given pathology in either an independent model (x-axis) or an adjusted model accounting for all other pathologies (y-axis). Each point represents a protein, color-coded by significance pattern: black for proteins significant in both models, red for those significant in the independent model only, and blue for those significant in the adjusted model only. The dashed diagonal line indicates equal effect sizes across models.

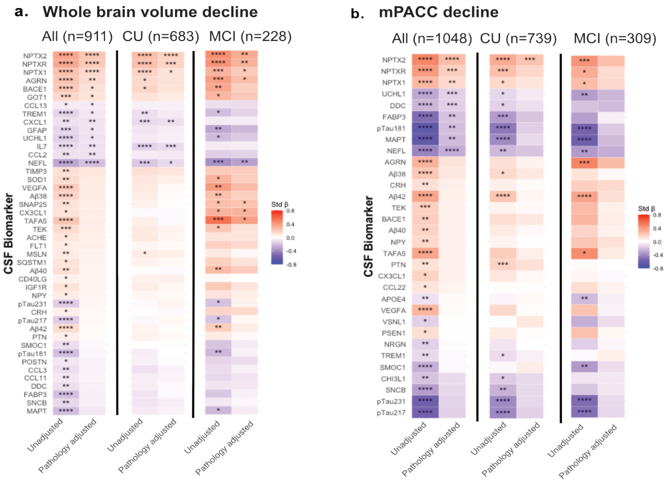


**Supplemental Figure 9: Prediction of whole brain volume atrophy and cognitive decline (mPACC v1) by DAPs in BioFINDER-2.**

**Panel a** shows heatmaps of standardized beta coefficients for the association between baseline DAP levels and annualized atrophy rate in whole brain volume, removing lateral ventricles volume and adjusted for ICV. **Panel b** presents analogous heatmaps for prediction of longitudinal change in modified Preclinical Alzheimer Cognitive Composite(mPACC) score. For each outcome, three heatmaps are displayed side-by-side: all participants, cognitively unimpaired (CU), and mild cognitive impairment (MCI). Within each heatmap, rows represent DAPs and the two columns correspond to the unadjusted protein association (left) and the association adjusted for baseline pathology burden (right). Cell color indicates direction (red for positive, blue for negative association) and strength (color intensity proportional to standardized beta value), with grey denoting non-significance. All models are adjusted for age, sex, and CSF protein levels, with additional adjustment for education in models of cognitive decline.*$P_{FDR} < 0.05$, **$P_{FDR} < 0.01$, ***$P_{FDR} < 0.001$, ****$P_{FDR} < 0.0001$.

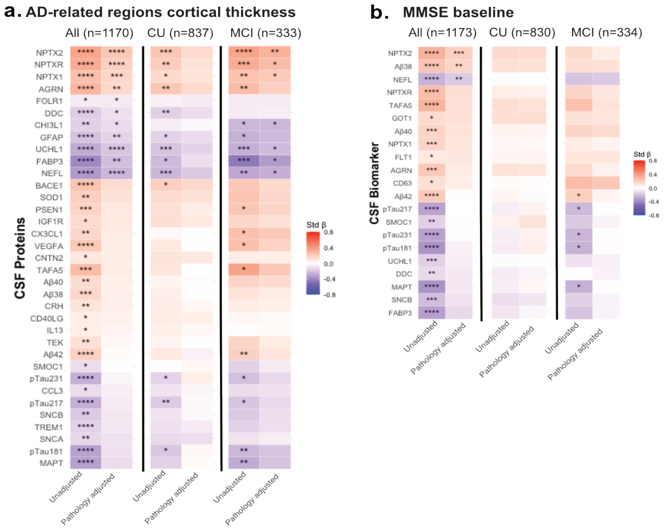


**Supplemental Figure 10: Prediction of cortical thickness in AD-specific regions and cognitive scores (MMSE) by DAPs in BioFINDER-2.**

**Panel a** shows heatmaps of standardized beta coefficients for the association between baseline DAP levels and cortical thickness in AD-signature regions. **Panel b** presents analogous heatmaps for prediction of scores in Mini-Mental State Examination (MMSE). For each outcome, three heatmaps are displayed side-by-side: all participants, cognitively unimpaired (CU), and mild cognitive impairment (MCI). Within each heatmap, rows represent DAPs and the two columns correspond to the unadjusted protein association (left) and the association adjusted for baseline pathology burden (right). Cell color indicates direction (red for positive, blue for negative association) and strength (color intensity proportional to standardized beta value), with grey denoting non-significance. All models are adjusted for age, sex, and CSF protein levels, with

| | All | Alzheimer's Disease Pathology | | α-Synuclein Pathology | | Vascular Pathology | |
|---|---|---|---|---|---|---|---|
| | | No | Yes | No | Yes | No | Yes |
| | N=1658 | 1290 | 368 | 1344 | 314 | 1112 | 546 |
| **Age at baseline (years)** | 72.2 (61.6 - 77.3) | 71.5 (59.1 - 77.1) | 73.9 (69.4 - 77.8) | 71.4 (59.7 - 77.0) | 74.4 (69.5 - 78.1) | 67.7 (56.8 - 75.4) | 76.2 (72.5 - 79.5) |
| **Sex- Female [N(%)]** | 841 (50.7%) | 676 (52.4%) | 165 (44.8%) | 636 (47.3%) | 205 (65.3%) | 523 (47.0%) | 318 (58.2%) |
| ***APOE* ε4 positivity [N (%)]** | 785 (47.4%) | 685 (53.1%) | 100 (27.2%) | 631 (47.0%) | 154 (49.0%) | 501 (45.1%) | 284 (52.0%) |
| **MMSE** | 28.0 (26.0 - 29.0) | 29.0 (27.0 - 30.0) | 24.0 (20.0 - 27.0) | 29.0 (26.0 - 30.0) | 27.0 (24.0 - 29.0) | 29.0 (27.0 - 30.0) | 27.0 (24.0 - 29.0) |
| **Cognitive Status – [N (%)]** | | | | | | | |
| *Cognitively Unimpaired (CU)* | 919 (55.4%) | 880 (68.2%) | 39 (10.6%) | 819 (60.9%) | 100 (31.9%) | 523 (47.0%) | 318 (58.2%) |
| *Mild Cognitive Impairment (MCI)* | 412 (24.9%) | 305 (23.6%) | 107 (29.1%) | 312 (23.2%) | 100 (31.9%) | 501 (45.1%) | 284 (52.0%) |
| *Dementia* | 327 (19.7%) | 105 (8.1%) | 222 (60.3%) | 213 (15.9%) | 114 (36.3%) | 723 (65.0%) | 196 (35.9%) |

additional adjustment for education in models of cognitive scores. *$P_{FDR}$ < 0.05, **$P_{FDR}$ < 0.01, ***$P_{FDR}$ < 0.001, ****$P_{FDR}$ < 0.0001.

**Supplemental Table 1: Demographics of included participants divided by each pathology**

Data shown as mean (95%CI) unless specified otherwise. Abbreviations: APOE, apolipoprotein E; CU, Cognitively Unimpaired; MCI, Mild cognitive impairment; MMSE, Mini-Mental State Examination; SUVR, standardized uptake value ratio.

| Participants (N) | No. 749 |
|---|---|
| **Age at baseline (years)** | 74.2 (68.0 – 78.0) |
| **Sex- Male [N (%)]** | 398 (53.1%) |
| ***APOE* ε4 positivity [N (%)]** | 360 (48.1%) |
| **MMSE** | 28.0 (26.0 - 29.0) |
| **Cognitive Status – [N (%)]** | |
| *Cognitively Unimpaired (CU)* | 385 (51.4%) |
| *Mild Cognitive Impairment (MCI)* | 198 (26.4%) |
| *Dementia* | 166 (22.2%) |
| **Alzheimer's Disease Pathology [N (%)] [a]** | 161 (21.5) |
| **α-Synuclein Pathology [N (%)] [b]** | 241 (32.2) |
| **Vascular Pathology [N (%)] [c]** | 241 (32.2) |

**Supplemental Table 2: Demographics of subcohort of participants with both CNS and Inflammation NULISA panel analyzed in CSF and CNS panel analyzed in plasma.**

Data shown as mean (95%CI) unless specified otherwise. Abbreviations: APOE, apolipoprotein E; CU, Cognitively Unimpaired; MCI, Mild cognitive impairment; MMSE, Mini-Mental State Examination; SUVR, standardized uptake value ratio.

a. Aβ positivity determined by [$^{18}$F]flutemetamol SUVR >1.033 (BioFINDER-2) or >1.138 (BioFINDER-1), or CSF Ab42/40 < 0.072 (Lumipulse G). Tau positivity defined by [$^{18}$F]RO948 tau-PET Braak I-IV SUVR > 1.362

b. Synuclein pathology positivity determined by positive α-synuclein Seed amplification assays (SAA).

c. WML positivity set at >0.59% of intracranial volume (ICV), top tertile

| Participants BF2 CU/MCI (N) | N=1173 |
|---|---|
| **Age at baseline (years)** | 70.6 (58.3 - 76.7) |
| **Sex- Female [N(%)]** | 575 (49.0%) |
| ***APOE* ε4 positivity [N (%)]** | 559 (47.7%) |
| **MMSE** | 29.0 (28.0 - 30.0) |
| **Cognitive Status – [N (%)]** | |
| *Cognitively Unimpaired (CU)* | 839 (71.5 %) |
| *Mild Cognitive Impairment (MCI)* | 334 (28.5%) |
| **Alzheimer's Disease Pathology [N(%)] [a]** | 136 (11.6%) |
| **α-Synuclein Pathology [N(%)] [b]** | 119 (10.1%) |
| **Vascular Pathology [N(%)] [c]** | 320 (27.3%) |

**Supplemental Table 3: Demographics of included BioFINDER-2 CU/MCI participants with continuous data available for pathologies**

Data shown as mean (95%CI) unless specified otherwise. Abbreviations: APOE, apolipoprotein E; CU, Cognitively Unimpaired; MCI, Mild cognitive impairment; MMSE, Mini-Mental State Examination; SUVR, standardized uptake value ratio.

a. Aβ positivity determined by [$^{18}$F]flutemetamol SUVR >1.033 (BioFINDER-2) or >1.138 (BioFINDER-1), or CSF Ab42/40 < 0.072 (Lumipulse G). Tau positivity defined by [$^{18}$F]RO948 tau-PET Braak I-IV SUVR > 1.362

b. Synuclein pathology positivity determined by positive α-synuclein Seed amplification assays (SAA).

c. WML positivity set at >0.59% of intracranial volume (ICV), top tertile

| Participants BF2 CU/MCI longitudinal (N) | N=656 |
|---|---|
| **Age at baseline (years)** | 67.9 (57.6 - 75.6) |
| **Sex- Female [N(%)]** | 328 (50.0%) |
| ***APOE* ε4 positivity [N (%)]** | 326 (49.7%) |
| **MMSE** | 29.0 (28.0 - 30.0) |
| **Cognitive Status – [N (%)]** | |
| *Cognitively Unimpaired (CU)* | 485 (73.9 %) |
| *Mild Cognitive Impairment (MCI)* | 171 (26.2%) |
| **Alzheimer's Disease Pathology [N(%)] [a]** | 78 (11.9%) |
| **α-Synuclein Pathology [N(%)] [b]** | 64 (9.8%) |
| **Vascular Pathology [N(%)] [c]** | 173 (26.4%) |

**Supplemental Table 4: Demographics of included BioFINDER-2 CU/MCI participants with longitudinal data available for pathologies**

Data shown as mean (95%CI) unless specified otherwise. Abbreviations: APOE, apolipoprotein E; CU, Cognitively Unimpaired; MCI, Mild cognitive impairment; MMSE, Mini-Mental State Examination; SUVR, standardized uptake value ratio.

a. Aβ positivity determined by [$^{18}$F]flutemetamol SUVR >1.033 (BioFINDER-2) or >1.138 (BioFINDER-1), or CSF Ab42/40 < 0.072 (Lumipulse G). Tau positivity defined by [$^{18}$F]RO948 tau-PET Braak I-IV SUVR > 1.362

b. Synuclein pathology positivity determined by positive α-synuclein Seed amplification assays (SAA).

c. WML positivity set at >0.59% of intracranial volume (ICV), top tertile
